# Comparative effectiveness of initiating nifedipine versus labetalol in early gestation for chronic hypertension in pregnancy

**DOI:** 10.1101/2025.08.05.25333078

**Authors:** Chase D. Latour, Madison Ponder, Kim Boggess, Jessie K. Edwards, Michele Jonsson Funk, Sharon Peacock Hinton, Mireille E. Schnitzer, Elizabeth A. Suarez, Mollie E. Wood

**Author notes:** **Corresponding Author:** Chase Latour, 170 Rosenau Hall, CB #7400, 135 Dauer Drive, Chapel Hill, NC 27599-7400.

## Abstract

**Background:** Limited data exist on the comparative effectiveness and safety of first-line medications to treat chronic hypertension in pregnancy. We compared nifedipine versus labetalol initiation before 23 weeks’ gestation for risk of pregnancy loss (miscarriage or stillbirth) and preterm birth (<37 weeks).

**Methods:** We identified pregnancies in Merative’s MarketScan commercial claims data with estimated last menstrual periods between July 1, 2016 and February 1, 2022. We included those with a new nifedipine or labetalol fill before 23 weeks’ gestation. We estimated risks, risk differences (RDs) per 100 pregnancies, and risk ratios (RRs) among nifedipine versus labetalol initiators using a weighted Aalen-Johansen estimator to account for confounding and informative censoring. We also conducted a *post hoc* analysis to estimate the comparative effect of nifedipine versus labetalol on risk of preeclampsia.

**Results:** We identified 987 nifedipine and 4,570 labetalol initiators during the study period. The weighted risk of pregnancy loss was 7.0% among nifedipine and 6.0% among labetalol initiators (RD=1.1 [-0.7, 2.8]; RR=1.18 [0.87, 1.59]). Weighted risks of preterm birth were 29.4% versus 22.2%, respectively (RD=7.2 [3.9, 10.5], RR=1.32 [1.13, 1.55]). In *post hoc* analyses, the weighted risks of preeclampsia were 38.6% versus 33.9%, respectively (RD=4.7 [0.7, 8.7], RR=1.14 [1.01, 1.28]). Findings were consistent across various sensitivity analyses.

**Conclusions:** Compared to labetalol initiators, nifedipine initiators experienced an increased risk of preterm birth but not pregnancy loss. The elevated risk of preterm birth may be explained by an increased risk of preeclampsia among nifedipine initiators, resulting in medically indicated preterm birth.

**Clinical Perspective:** *What is new?:*

- Initiating nifedipine, compared to labetalol, before 23 weeks’ gestation as a first-line antihypertensive was associated with a 7.2 percentage point increased risk of preterm live birth (95% CI: 3.9, 10.5) and 4.7 percentage point increased risk of preeclampsia (95% CI: 0.7, 8.7).
- Nifedipine and labetalol were associated with a similar risk of pregnancy loss (risk difference: 1.1 per 100 pregnancies [95% CI: -0.7, 2.8]).
- These findings were robust across multiple sensitivity analyses.

*What are the clinical implications?:*

- Initiating labetalol, compared to nifedipine, prior to 23 weeks’ gestation for treatment of chronic hypertension in pregnancy may lead to lower risk of preeclampsia and preterm live birth in real-world settings.
- While high-quality data from clinical trials clearly support antihypertensive therapy for chronic hypertension in pregnancy, our findings illustrate that the first-line medications may have different comparative safety and effectiveness profiles.
- These data are important for informing shared decision-making between providers and patients managing chronic hypertension in pregnancy.

## Introduction

Chronic hypertension in pregnancy—hypertension that is identified or diagnosed prior to 20 weeks’ gestation^1^—is associated with elevated risks of adverse maternal and neonatal outcomes such as medically indicated preterm birth and perinatal death.^1,2^ Given chronic hypertension’s increasing prevalence to ∼4% among deliveries in the United States (US),^3^ it is critical to identify interventions that improve perinatal outcomes.

A major advance in care came from the Chronic Hypertension and Pregnancy (CHAP) trial—a multi-center, open-label trial—which found that pregnant persons randomized to stricter blood pressure control (<140/90 mmHg), compared to less-strict control (<160/105 mmHg), via antihypertensive pharmacotherapy had improved pregnancy outcomes without an increased risk of small-for-gestational age infants.^4^ These findings led to immediate changes in clinical guidelines, with societies now recommending pharmacologic treatment of both mild and severe chronic hypertension during pregnancy.^5,6^

While the CHAP trial provided strong evidence to support the use of antihypertensives for chronic hypertension in pregnancy, questions remain as to which first-line antihypertensive— nifedipine or labetalol—has a more favorable safety and effectiveness profile. A recent meta-analysis of low-to-moderate quality trials suggested labetalol may reduce the risk of preeclampsia and preterm birth compared to nifedipine,^7^ but these findings were not supported by a secondary reanalysis of the CHAP trial.^8^ A large trial is ongoing in the United Kingdom to compare these drugs among pregnant people with chronic or pregnancy-induced hypertension,^9^ but its findings may have limited generalizability (e.g., different adherence patterns among trial participants versus real-world patients).^10–12^ Observational studies offer an important means of quantifying these comparative effects in real-world populations,^11^ but prior work is subject to important sources of bias—for example, restricting to pregnancies that ended in delivery.^13–16^ These biases make it difficult to determine whether differences from trial findings reflect true population differences or are artifacts of study design.^17,18^

Target trial emulation is an increasingly common framework for minimizing bias in observational studies. It involves designing and analyzing data to emulate (mirror) a hypothetical randomized trial—the “target trial”.^19–22^ In this study, we emulated a target trial to estimate the effect of initiating nifedipine versus labetalol before 23 weeks of gestation on the risk of pregnancy loss and preterm live birth using commercial insurance claims data. To avoid selection bias, we included pregnancies with unobserved outcomes and employed sensitivity analyses to assess varying assumptions about these pregnancies.

## Methods

Our hypothetical target trial and its emulation is shown in Table 1 and described below.

**Table 1.**
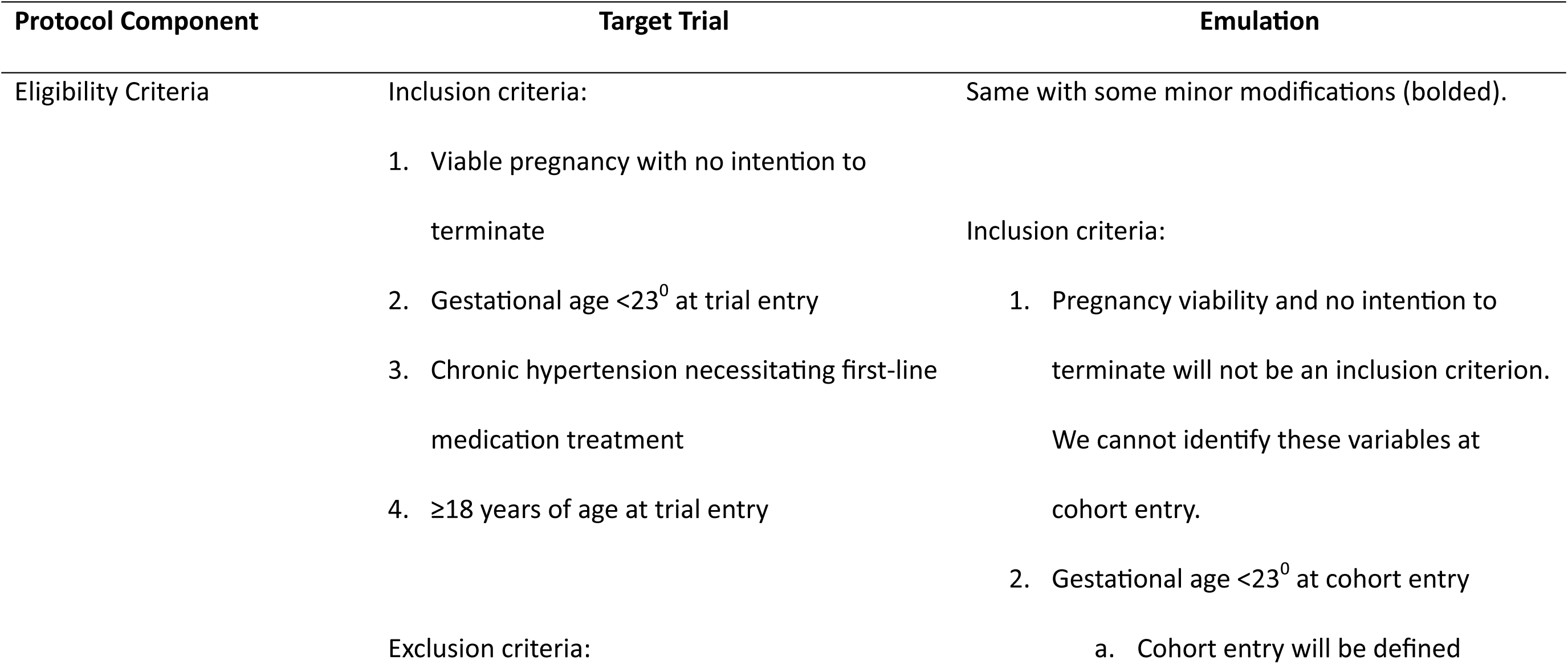
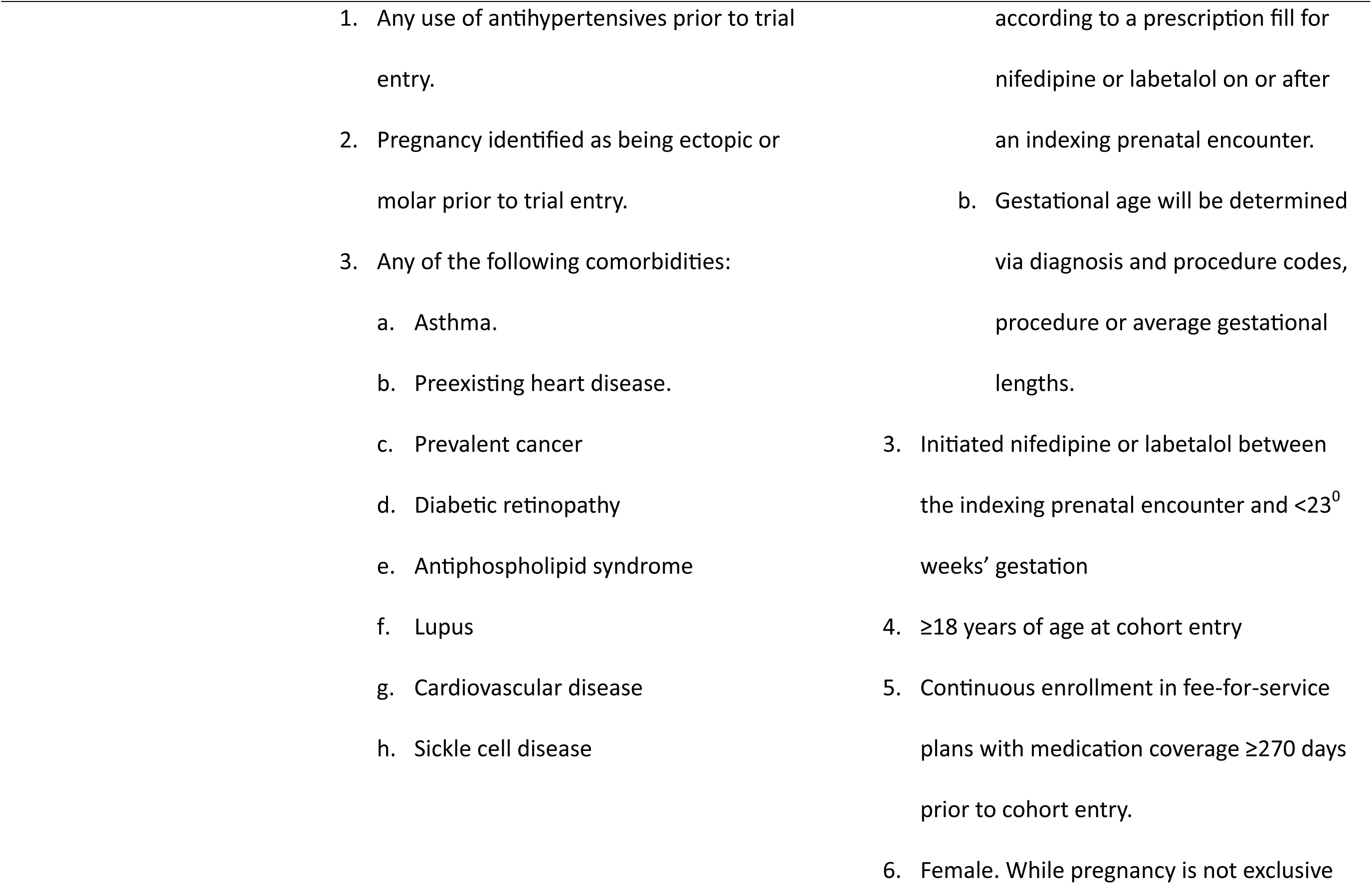
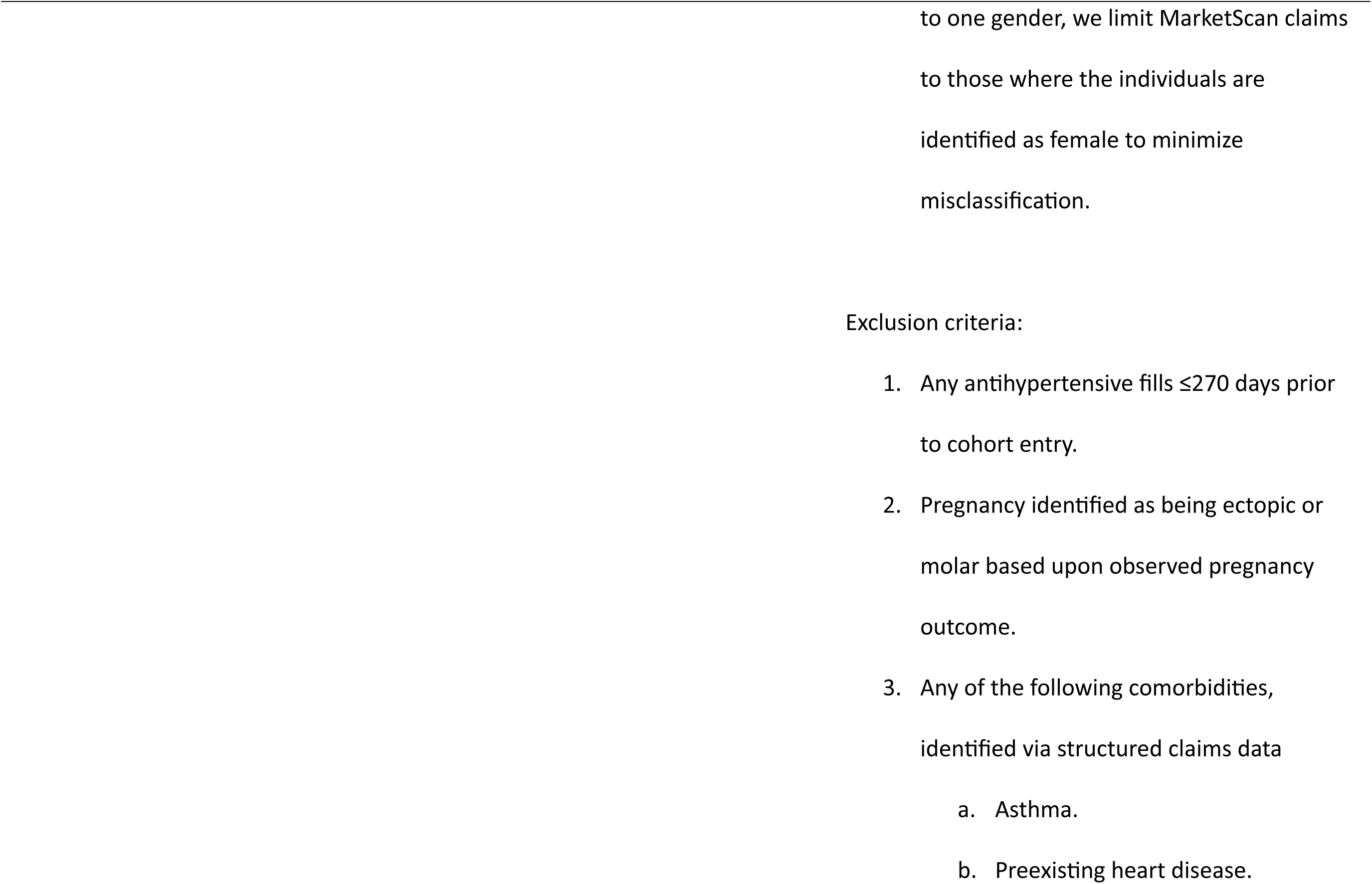
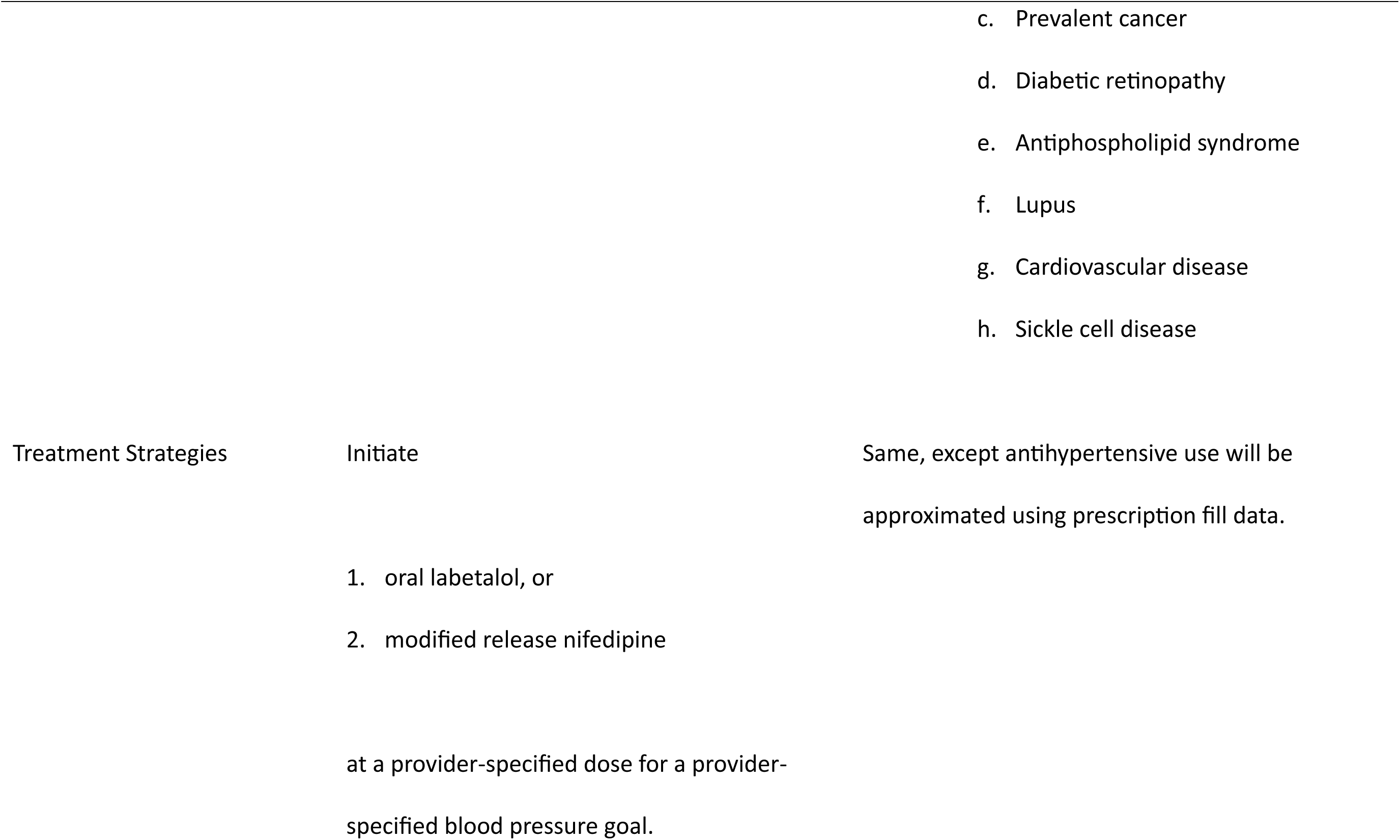
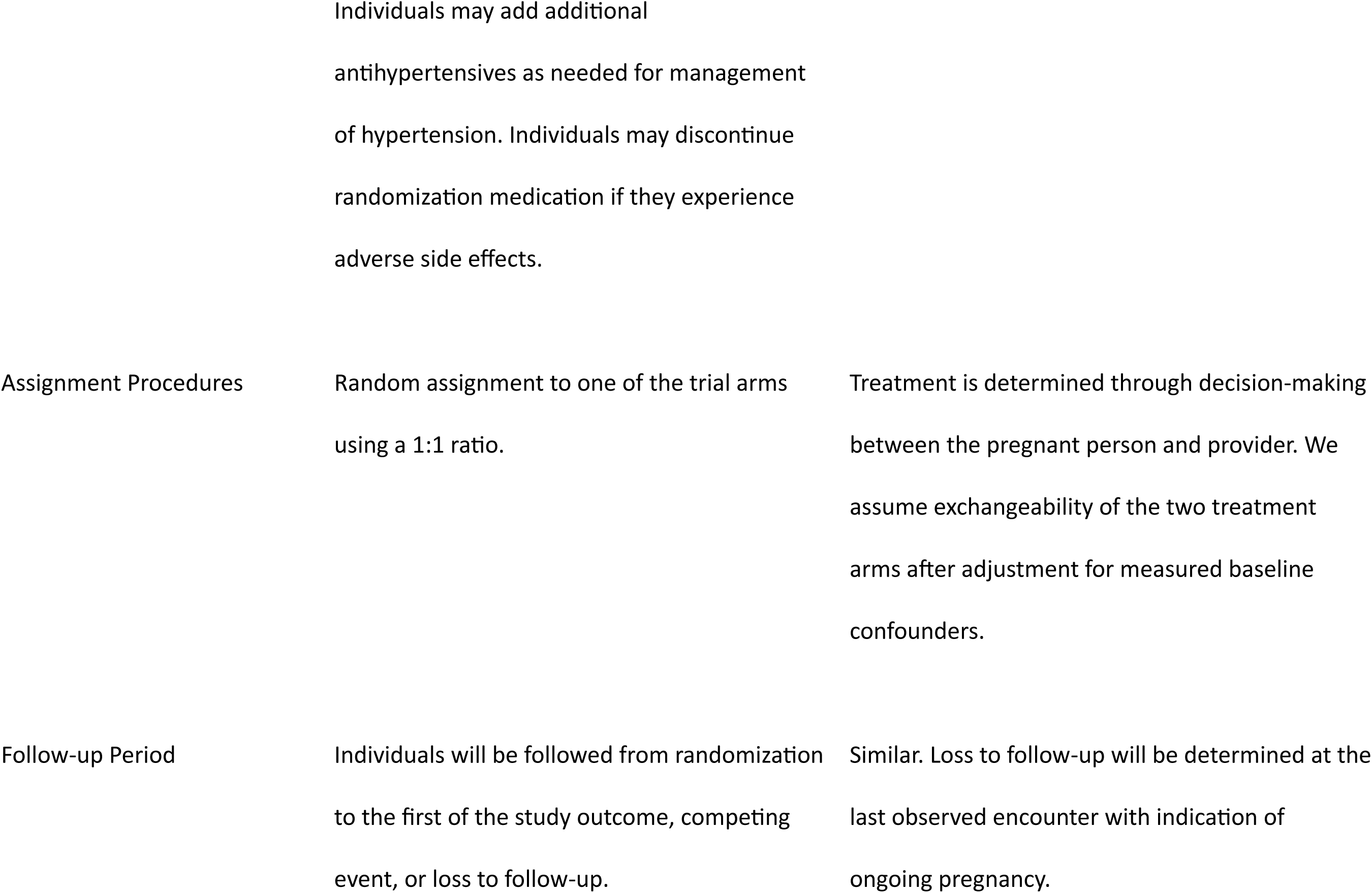
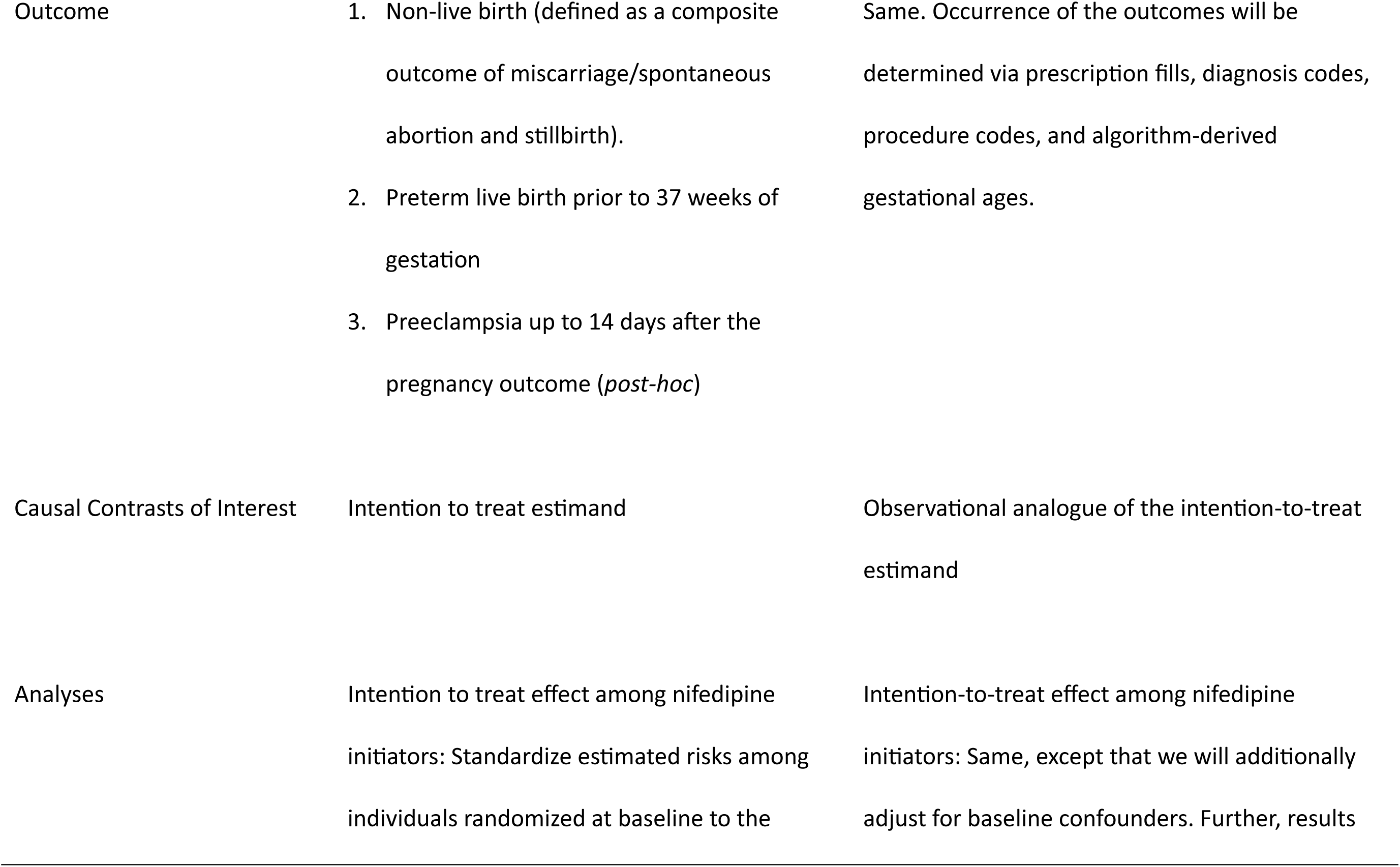
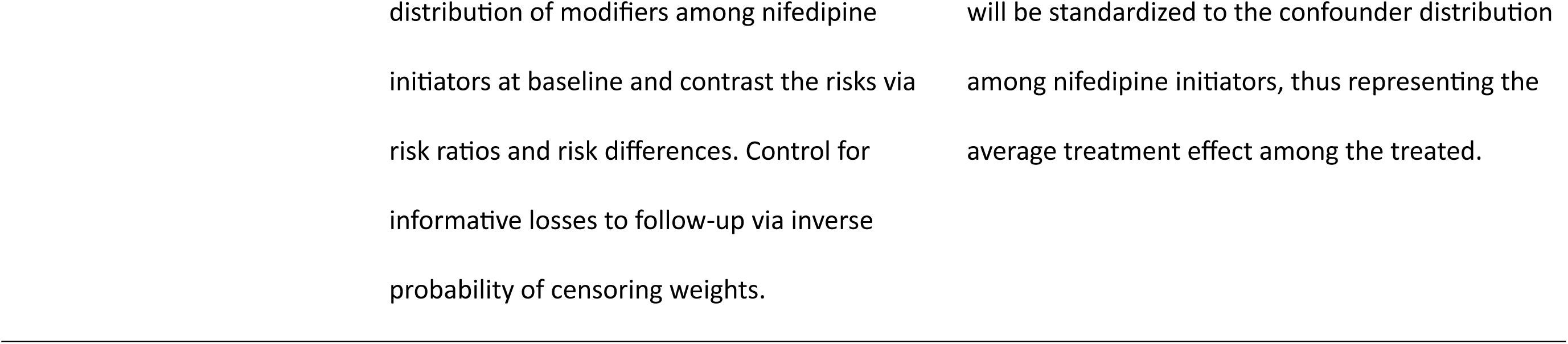
Protocol components for our target trial and its emulation in the Merative MarketScan Commercial Claims and Encounters Database.

### Target Trial

We designed a hypothetical target trial to compare initiation of nifedipine or labetalol at <23 weeks and 0 days (<23^0^ weeks) of gestation for chronic hypertension.^4^ Pregnant patients would be eligible for the target trial if they were diagnosed with chronic hypertension necessitating first-line antihypertensive pharmacotherapy, had a viable pregnancy before 23^0^ weeks’ gestation with no intention to terminate, no prior antihypertensive use, and were ≥18 years of age (full eligibility criteria shown in Table 1). In this hypothetical target trial, participants would be randomized to initiate modified-release nifedipine or oral labetalol to achieve a provider-specified blood pressure goal with ability to add or switch antihypertensives as needed.

### Emulation

#### Data

Data for this observational study came from the Merative MarketScan Commercial Claims and Encounters Database (hereafter “MarketScan”), which contains adjudicated outpatient, inpatient, and prescription claims from enrollees of employer-sponsored health insurance in the US. Outpatient and inpatient claims contain International Classification of Diseases, 9^th^ and 10^th^ edition, diagnosis and procedure codes; Current Procedural Terminology codes; and Healthcare Common Procedure Coding System codes. Prescription claims are limited to outpatient prescription fills. The study was approved by the Institutional Review Board at the University of North Carolina at Chapel Hill (#22-2471).

#### Pregnancies

A fundamental principle of target trial emulation is that (1) the start of follow-up (time zero) should align with the time when treatment is assigned, and (2) only information available before time zero should be used to determine study eligibility. However, this principle can be challenging to uphold in pregnancy studies using administrative data because pregnancies are typically identified based on observed outcomes such as delivery.^23,24^ In studies of prenatal medication use, relying on such outcome-based pregnancy identification strategies violates this principle because study entry then requires an observed pregnancy outcome, which, by definition, occurs after treatment assignment. Further, pregnancies with unobserved outcomes are likely disproportionately comprised of early losses and induced abortions, which are less often recorded in administrative claims data; thus, failure to include these pregnancies may induce bias.^13–16^

We used a novel algorithm adapted from prior work to identify a cohort of pregnancies, regardless of outcome, in MarketScan data, to allow for valid emulation of study entry in the hypothetical target trial (Methods S1-S2).^25–34^ Pregnancies with unobserved outcomes were considered lost to follow-up at their last claim with evidence of ongoing pregnancy. For pregnancies with unobserved outcomes and no gestational age information, we assumed a gestational age of 9^0^ weeks at their first claim with evidence of ongoing pregnancy, which corresponds to the US mean gestational age at prenatal care initiation.^35^ We limited our analyses to those pregnancies with estimated last menstrual periods (LMPs) between July 1, 2016 and February 1, 2022.

#### Eligibility Criteria

We identified pregnancies with ≥1 fill for modified-release nifedipine or oral labetalol between their indexing prenatal encounter and 22^6^ weeks’ gestation, excluding those with antihypertensive fills in the prior 270 days (i.e., those on antihypertensives prior to pregnancy) (Figure S1, Table S1, Supplemental File 2).^36^ The first qualifying fill defined initiation and the index date. The primary analysis did not use chronic hypertension diagnosis codes to identify patients because chronic hypertension is the primary reason for early gestation initiation of nifedipine or labetalol and is likely under-coded.^1,37^ We excluded pregnant patients without continuous medical and medication coverage for 270 days pre-index (31-day allowable gap); <18 years at the index; with an ectopic or molar pregnancy outcome; or with preexisting asthma, heart disease, prevalent cancer, diabetic retinopathy, antiphospholipid syndrome, lupus, cardiovascular disease, or sickle cell disease using diagnosis and procedure codes and medications recorded in claims 270 days pre-index (Table S2). For patients with multiple pregnancies, only the first qualifying pregnancy was included.

#### Treatment Strategies

We assumed nifedipine and labetalol were initiated to achieve a provider- and patient-determined blood pressure goal. Before CHAP’s publication (April 2, 2022), the recommended target blood pressure for chronic hypertension among pregnant people was <160/110 mmHg. After CHAP, the target changed to <140/90 mmHg.^4,5^ This shift should only affect only those who initiated nifedipine or labetalol between CHAP’s publication and July 12, 2022 (23 weeks after February 1, 2022).

#### Treatment Assignment

Treatment was not randomized; instead, we identified a set of potential confounders using a directed acyclic graph (DAG) (Figure S2).^38,39^ Measured potential confounders included gestational age; maternal age; year; substance use/abuse; nausea/vomiting; diabetes and antidiabetic prescription fills; migraine and triptan fills; preeclampsia; history of recurrent pregnancy loss; chronic health conditions (chronic kidney disease, obesity, thyroid disease and medication fills, mental health conditions and psychotropic medication fills, and hyperlipidemia); and healthcare use (number of prior outpatient prenatal encounters, number of prior inpatient admissions) (Table S3).

Most covariates were identified using claims ≤270 days before or ≤30 days after the index date (before the pregnancy outcome/loss to follow-up date) (Figure S1).^24^ Variables only assessed ≤270 days before the index date included substance use/abuse, nausea/vomiting, preeclampsia, migraine, recurrent pregnancy loss, teratogenic medication use, and healthcare use. Diagnosis and procedure codes and prescription fills used to identify variables are in Supplemental File 2.

#### Follow-up Period

Pregnancies were followed from treatment initiation (time zero) until the first instance of the study outcome, competing event (defined in *Primary Analyses*), or last claim with indication of ongoing pregnancy (i.e., loss to follow-up). We used a DAG to identify measured potential causes of loss to follow up (Figure S3),^40,41^ which included rurality and confounders.

#### Study Outcomes

We defined pregnancy loss as a composite outcome of miscarriage, unspecified abortion at any gestational age, and stillbirth. We defined preterm live birth as a composite outcome of singleton live birth, multiple live birth, or uncategorized delivery before 37 weeks’ gestation.

#### Primary Analyses

We used an active comparator new user cohort design to emulate the target trial.^42^ The target estimand was the observational analogue of the intention to treat effect in the nifedipine-initiating population. We estimated risks and treatment effects stratified by gestational age at initiation and then combined these estimates to derive risks in the overall population.

To address confounding, we estimated propensity scores (PSs) within stratum of gestational age at initiation (<14^0^ versus ≥14^0^ weeks) using logistic regression to predict treatment with nifedipine versus labetalol conditional upon measured confounders. Individuals with PSs in non-overlapping regions were excluded, and the logistic regression was re-fitted.^43,44^ The final PS model was selected based on convergence and standardized mean differences <0.1 within covariate strata (Methods S3). To standardize results to the group receiving nifedipine, we used treatment weights of 1 for nifedipine and PS/(1-PS) for labetalol initiators.^44^

We used stabilized inverse probability of censoring weights to address informative censoring.^41^ Within gestational age and treatment strata, we fit pooled logistic regression models to estimate the probability of remaining uncensored at timepoints corresponding to quintiles of follow-up dependent upon measured variables at baseline (Methods S4). Stabilized time-varying weights were calculated as the proportion remaining uncensored through the timepoint divided by the model-predicted probability of remaining uncensored at that time conditional on baseline covariates.

To estimate risks stratified by gestational age at initiation, we used a treatment- and censoring-weighted Aalen-Johansen estimator to calculate the risk of pregnancy loss and preterm birth within each gestational age stratum while accounting for competing events.^45^ For pregnancy loss, competing events included induced abortion and live birth;^46,47^ for preterm birth, they were induced abortion, pregnancy loss, and survival to 37 weeks’ gestation. We contrasted stratum-specific weighted risks across treatment groups as risk differences (RDs) per 100 pregnancies and risk ratios (RRs).

To estimate treatment effects in the overall population, we standardized the treatment- and censoring-weighted risks to the gestational age distribution at initiation among all nifedipine initiators. This approach preserved the confounding and censoring adjustment while producing marginal risk estimates representative of the treated population. We contrasted these risks using RDs and RRs and calculated Wald-type 95% confidence intervals (CIs) via bootstrap with 1,000 replicates (Methods S5).^48^

### Secondary Analyses

To evaluate whether the treatment effect varied for early versus later study outcomes, we estimated the risk of preterm birth at <28, <32, and <34 weeks’ gestation among the full cohort. To report risks of pregnancy loss at different gestational age cutoffs, we fit three Aalen-Johansen estimators to estimate the cumulative incidence at <12, 12-19, and ≥20 weeks’ gestation. These latter analyses were conducted among initiators before 12 weeks’ gestation, treating losses at prior gestational age intervals as competing events (e.g., losses at <12 weeks’ gestation were competing events to losses at 12-19 weeks).

### Post Hoc Analysis

The World Health Organization has expressed concern that calcium channel blockers, such as nifedipine, may increase the risk of preeclampsia in pregnancy,^49^ which, in turn, may increase risk of indicated preterm birth.^1,50^ After observing the results for preterm birth, we conducted a post hoc analysis to estimate the effect of nifedipine versus labetalol initiation before 23 weeks’ gestation on risk of preeclampsia. We thus identified a cohort of pregnancies without prevalent preeclampsia at their index date (i.e., no pre-index diagnosis codes).^46^ We defined preeclampsia ≤14 days post-pregnancy outcome according to (1) the first diagnosis code at any location of service and (2) the first inpatient diagnosis code (Methods S6).^51,52^ We then applied the methods from the primary analyses, additionally censoring pregnancies at their first insurance disenrollment post-index. Competing events included induced abortion, pregnancy loss, or not experiencing preeclampsia within 14 days after the pregnancy outcome.^47^

### Sensitivity Analyses

Primary analyses used censoring weights to account for informative loss to follow-up, assuming that censored outcomes were missing at random given measured covariates. However, some pregnancies’ outcomes may be missing not random (MNAR)—that is, missing because of the true, unobserved outcome (e.g., a miscarriage managed at home or an induced abortion not billed to insurance). To quantify potential bias if all unobserved outcomes were MNAR, we estimated full bounds on the RDs and RRs (sensitivity analysis 1 in Table 2).^53,54^ Specifically, we repeated the primary analysis for pregnancy loss under four assumptions about pregnancies with unobserved outcomes: 1) none had non-live births, 2) all had non-live births, 3) nifedipine initiators had non-live births and labetalol initiators had term live births, and 4) vice-versa. We applied the same approach to preterm live birth and preeclampsia (Methods S7).

**Table 2.**
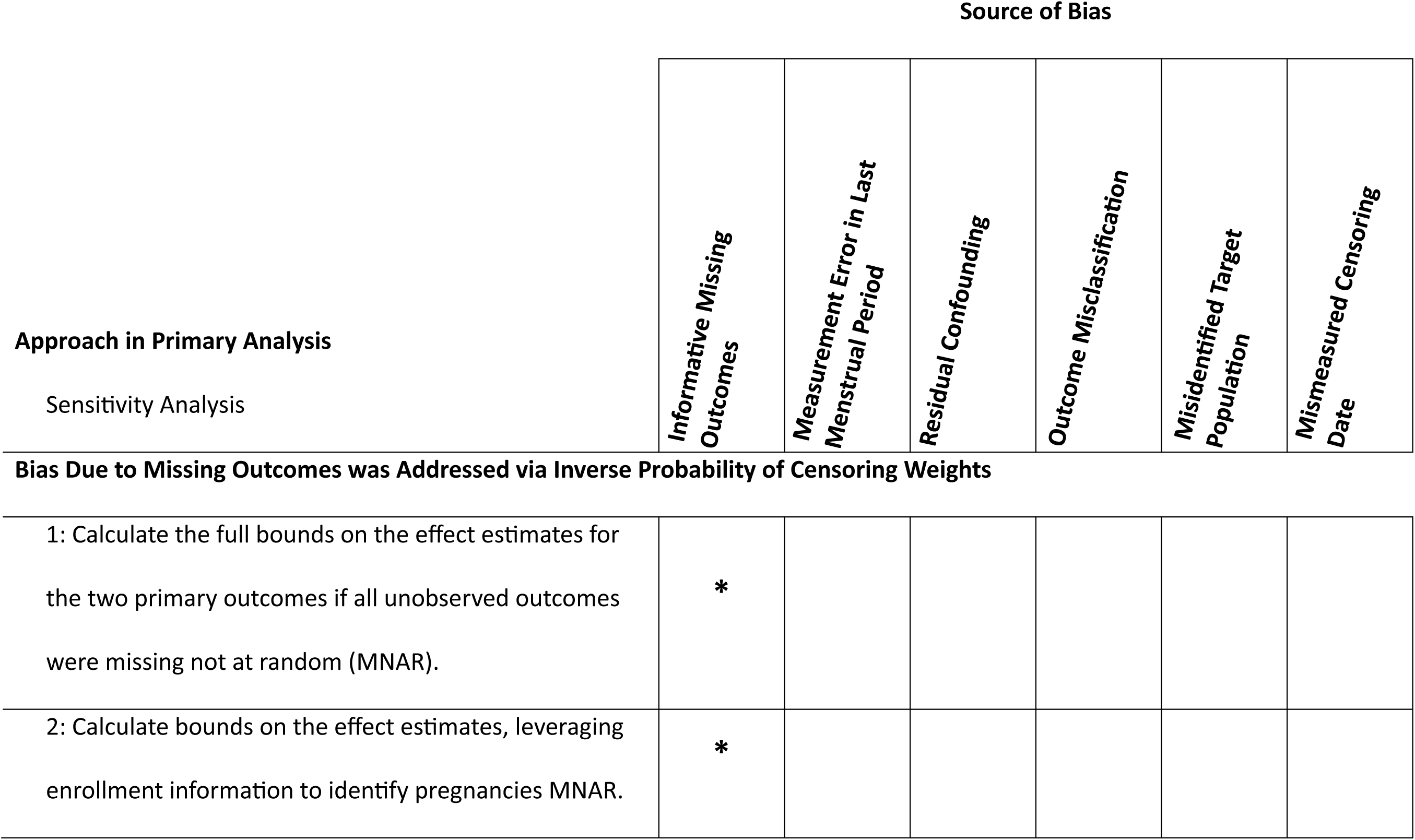
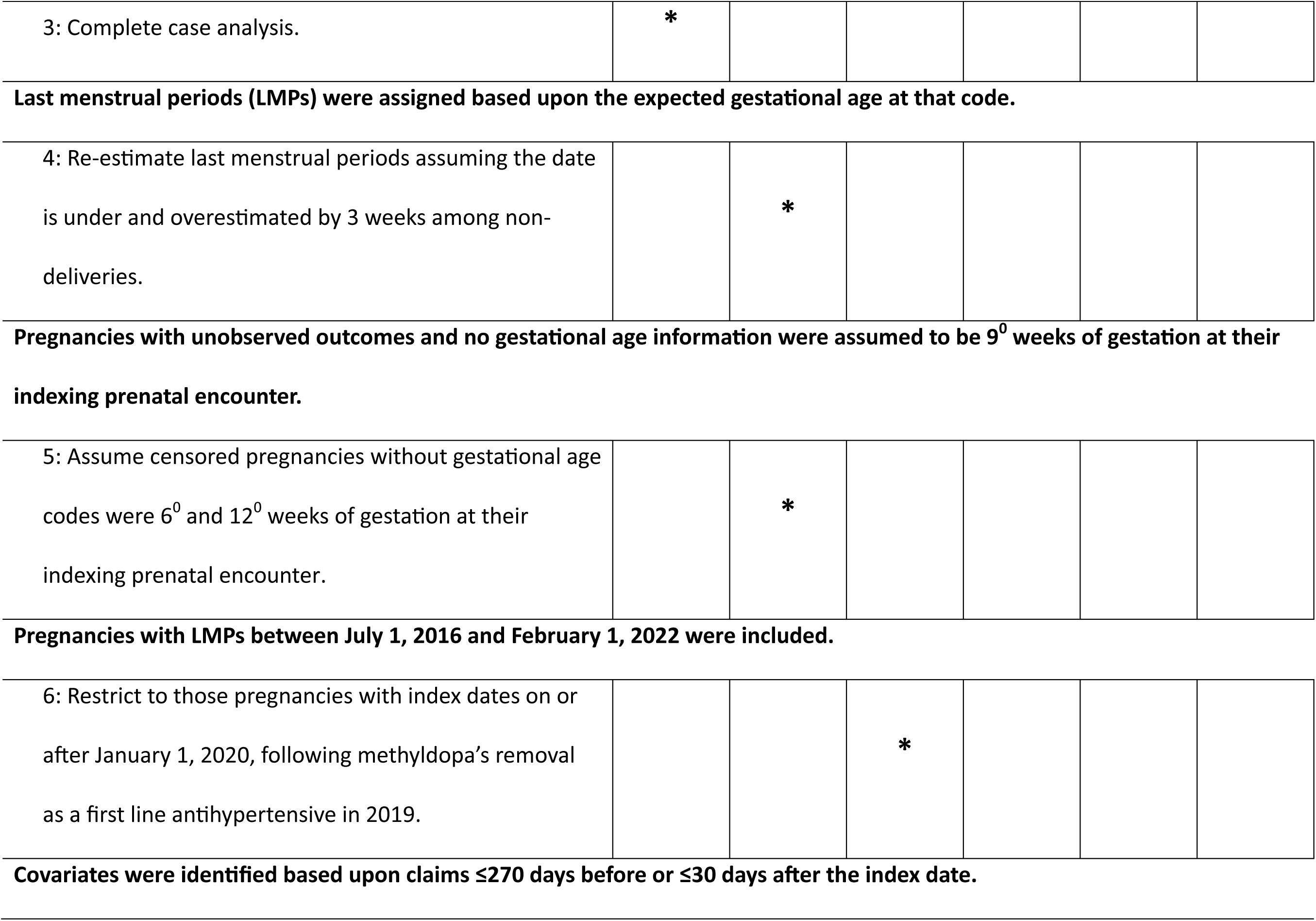
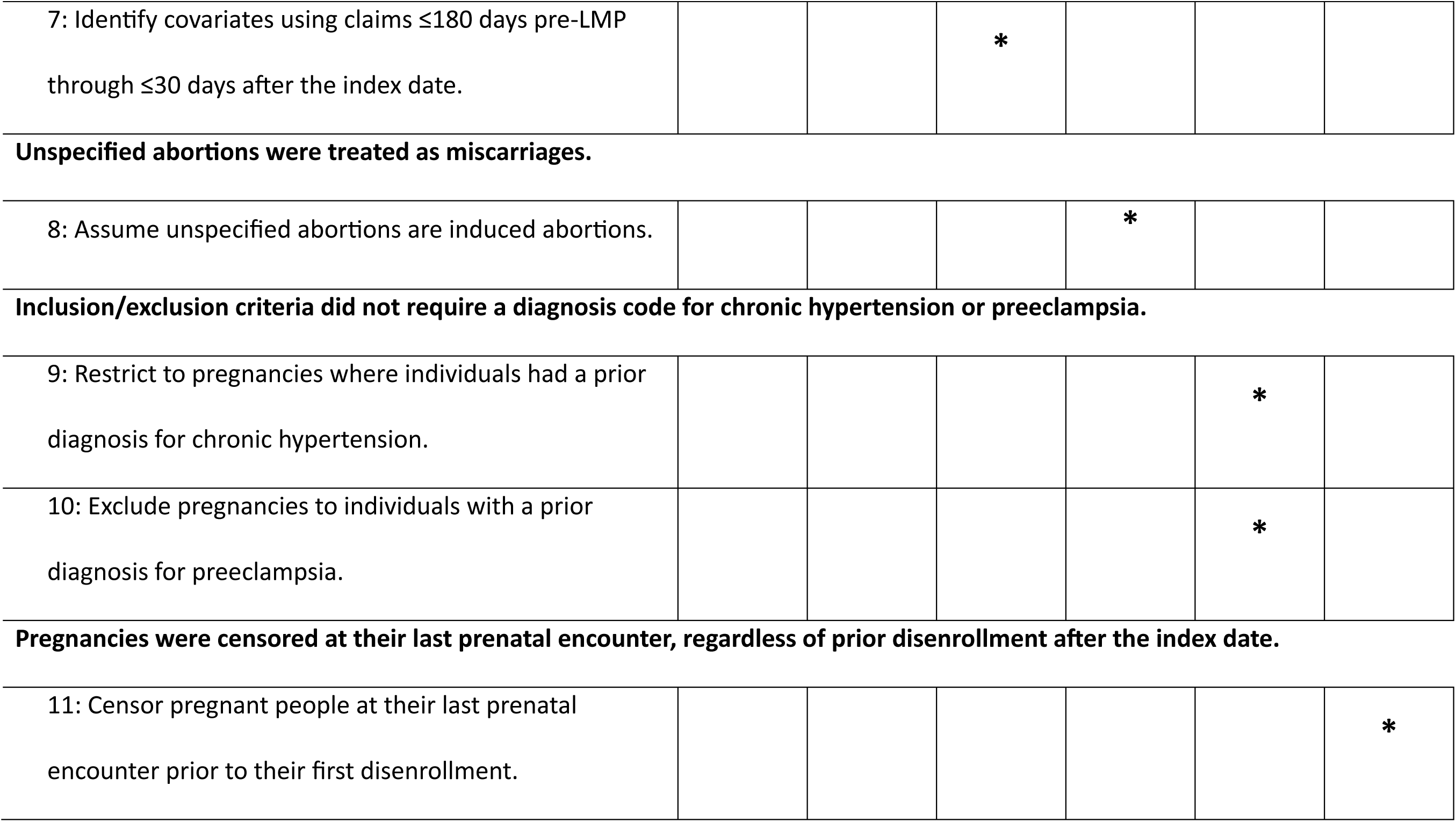
Sensitivity analyses and the sources of bias that they intended to address.

|  |  | Source of Bias |  |  |  |  |  |
| --- | --- | --- | --- | --- | --- | --- | --- |
| Approach in Primary Analysis |  | *Informative Missing Outcomes* | *Measurement Error in Last Menstrual Period* | *Residual Confounding* | *Outcome Misclassification* | *Misidentified Target Population* | *Mismeasured Censoring Date* |
| Sensitivity Analysis |  |  |  |  |  |  |  |
| Bias Due to Missing Outcomes was Addressed via Inverse Probability of Censoring Weights |  |  |  |  |  |  |  |
| 1: Calculate the full bounds on the effect estimates for the two primary outcomes if all unobserved outcomes were missing not at random (MNAR). |  | * |  |  |  |  |  |
| 2: Calculate bounds on the effect estimates, leveraging enrollment information to identify pregnancies MNAR. |  | * |  |  |  |  |  |

|  |  |
| --- | --- |
| 3: Complete case analysis. | * |

**Last menstrual periods (LMPs) were assigned based upon the expected gestational age at that code.**
|  |  |  |
| --- | --- | --- |
| 4: Re-estimate last menstrual periods assuming the date is under and overestimated by 3 weeks among non-deliveries. |  | * |

|  |  |  |
| --- | --- | --- |
| 5: Assume censored pregnancies without gestational age codes were 6<sup>0</sup> and 12<sup>0</sup> weeks of gestation at their indexing prenatal encounter. |  | * |

**Pregnancies with LMPs between July 1, 2016 and February 1, 2022 were included.**
|  |  |  |  |
| --- | --- | --- | --- |
| 6: Restrict to those pregnancies with index dates on or after January 1, 2020, following methyldopa's removal as a first line antihypertensive in 2019. |  |  | * |

**Covariates were identified based upon claims $\leq 270$ days before or $\leq 30$ days after the index date.**
|  |  |  |  |  |  |  |
| --- | --- | --- | --- | --- | --- | --- |
| 7: Identify covariates using claims $\leq 180$ days pre-LMP through $\leq 30$ days after the index date. | | | * | | | |

**Unspecified abortions were treated as miscarriages.**
|  |  |  |  |  |
| --- | --- | --- | --- | --- |
| 8: Assume unspecified abortions are induced abortions. |  |  |  | * |

**Inclusion/exclusion criteria did not require a diagnosis code for chronic hypertension or preeclampsia.**
|  |  |  |  |  |  |
| --- | --- | --- | --- | --- | --- |
| 9: Restrict to pregnancies where individuals had a prior diagnosis for chronic hypertension. |  |  |  |  | * |
| 10: Exclude pregnancies to individuals with a prior diagnosis for preeclampsia. |  |  |  |  | * |

**Pregnancies were censored at their last prenatal encounter, regardless of prior disenrollment after the index date.**
|  |  |  |  |  |  |  |
| --- | --- | --- | --- | --- | --- | --- |
| 11: Censor pregnant people at their last prenatal encounter prior to their first disenrollment. |  |  |  |  |  | * |

The width of the full bounds depends on the number of pregnancies assumed to be MNAR. To construct narrower bounds, we used MarketScan enrollment data to make assumptions about which pregnancies with unobserved outcomes were missing because of their true, unobserved pregnancy outcome (i.e., MNAR) versus missing for other, measured reasons (sensitivity analysis 2 in Table 2, Methods S9). We classified pregnancies with unobserved outcomes into two groups: (1) MNAR, or those with continued enrollment >31 days after loss to follow-up, and (2) missing at random given observed covariates, or those with disenrollment ≤31 days after loss to follow-up. We then repeated the primary analysis under four bounding assumptions for MNAR pregnancies: 1) all experienced the study outcome at risk period start, 2) none experienced a study outcome by risk period end, 3) only nifedipine initiators experienced the study outcome at risk period start, and 4) vice-versa (Methods S8). We assumed that informative censoring among pregnancies in group (2) was addressed via censoring weights. For preterm birth, we additionally constructed bounds under analogous assumptions using immediate pregnancy loss and term delivery as extremes.

We conducted a complete case analysis among pregnancies with observed outcomes, which assumed that outcomes were missing completely at random (sensitivity analysis 3 in Table 2). While this assumption is not supported by our DAG, which assumes multiple potential causes of missingness, it is possible that the bias from measurement error in gestational age and censoring date among pregnancies with unobserved outcomes outweighs bias due to missingness.^23,55^

Finally, we conducted a series of analyses to assess the robustness of our primary results to other assumptions including gestational age misestimation, residual confounding, outcome misclassification, incorrectly identifying the target population, and measurement error in censoring dates (sensitivity analyses 4-5, 6-7, 8, and 9-11, respectively, in Table 2, Methods S10).

### Descriptive Adherence Analyses

We described the percentage of initiators that (1) initiated an antihypertensive other than their initial exposure prior to their outcome or loss to follow-up and (2) experienced a >30-day gap in days’ supply of their initiation medication prior to the pregnancy outcome or loss to follow-up. We did not have sufficiently detailed covariate information to conduct a well-controlled per-protocol analysis.^56^ Instead, as a *post hoc* analysis, we constructed bounds on the per-protocol effect (Methods S11).^57^

Analyses were conducted using SAS Statistical Software (Cary, NC, USA). Figures were created using R Statistical Software (Vienna, Austria).

## Results

We identified 987 nifedipine and 4,570 labetalol initiators, with 49.3% and 52.0% of nifedipine and labetalol users initiating before gestational week 14, respectively (Table 3, Table S4, Figure S5). In the unweighted population, nifedipine initiation was more common after 2019 and nifedipine initiators were more likely to have ≥1 fill for an antidiabetic. We achieved sufficient balance on measured confounders via treatment weighting (Figure S6, Tables S4-S5).

**Table 3.**
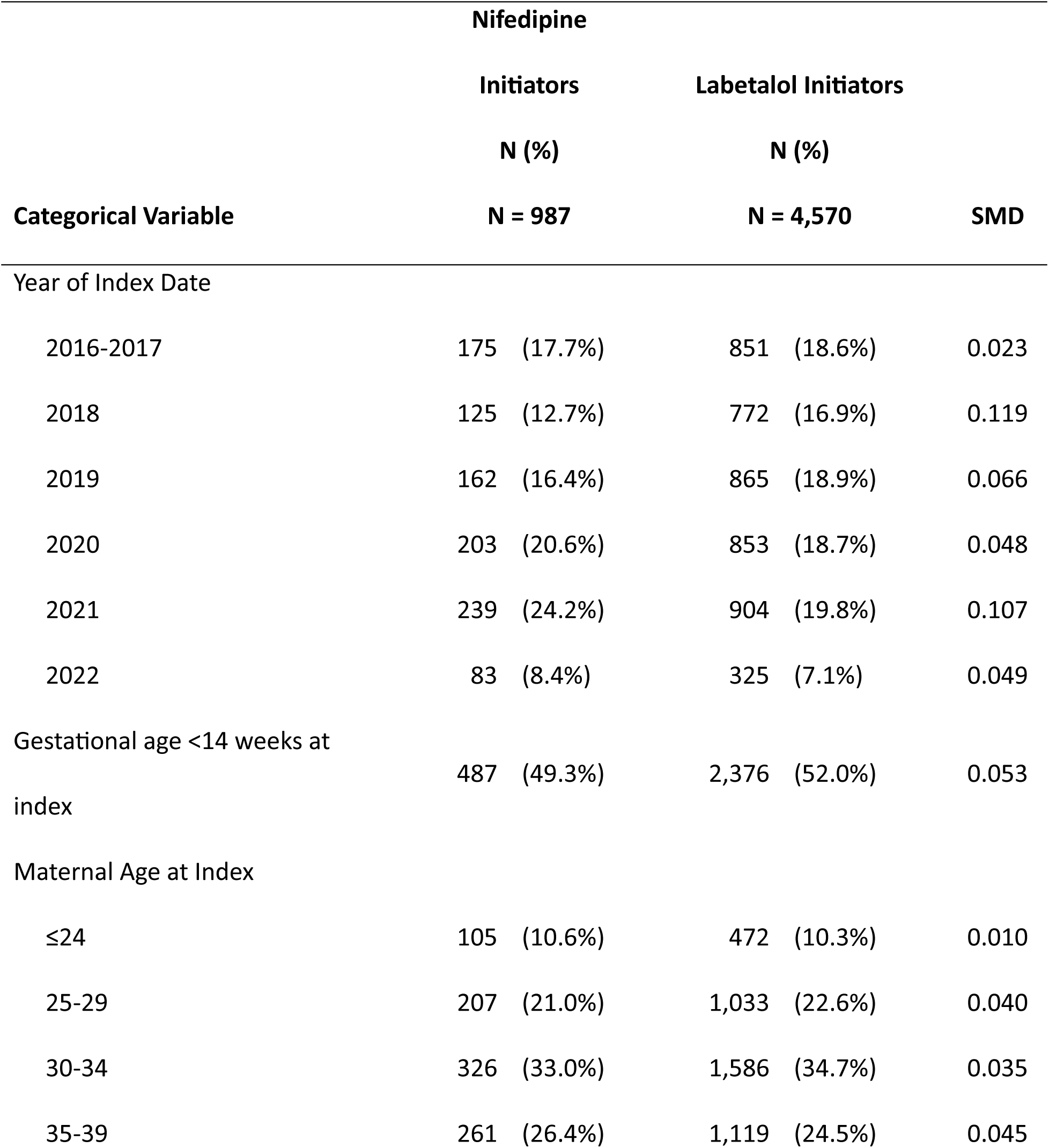
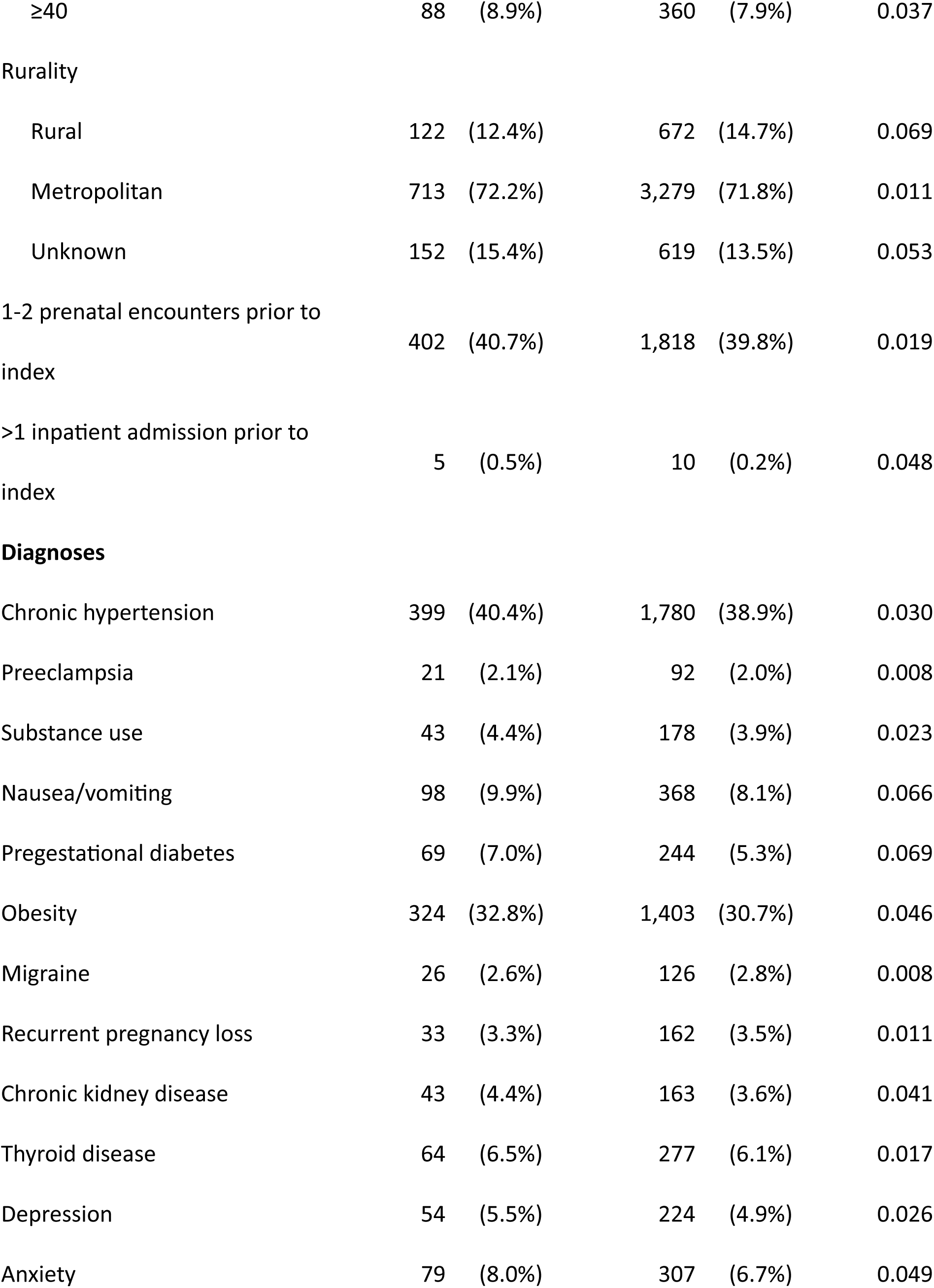
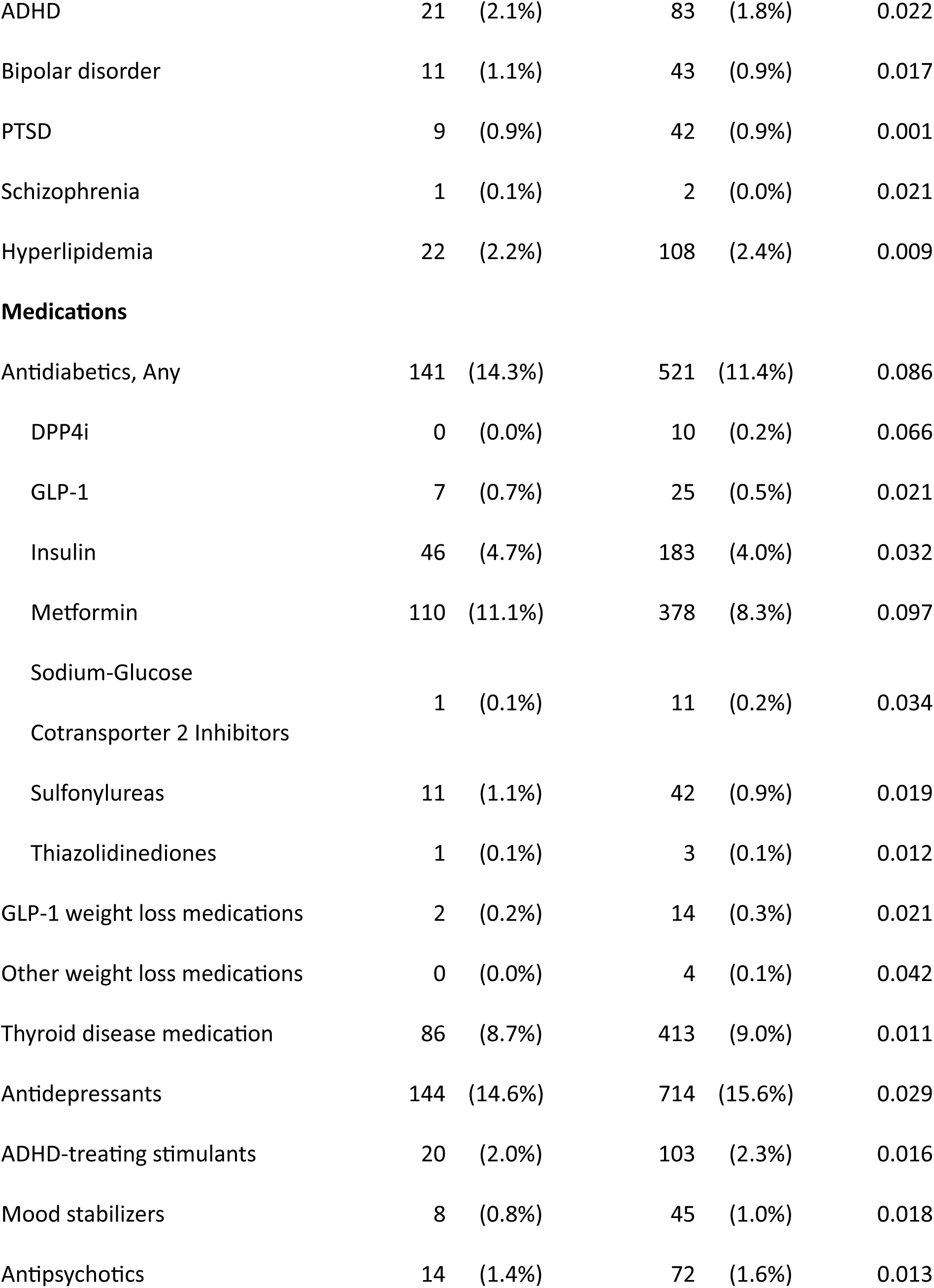
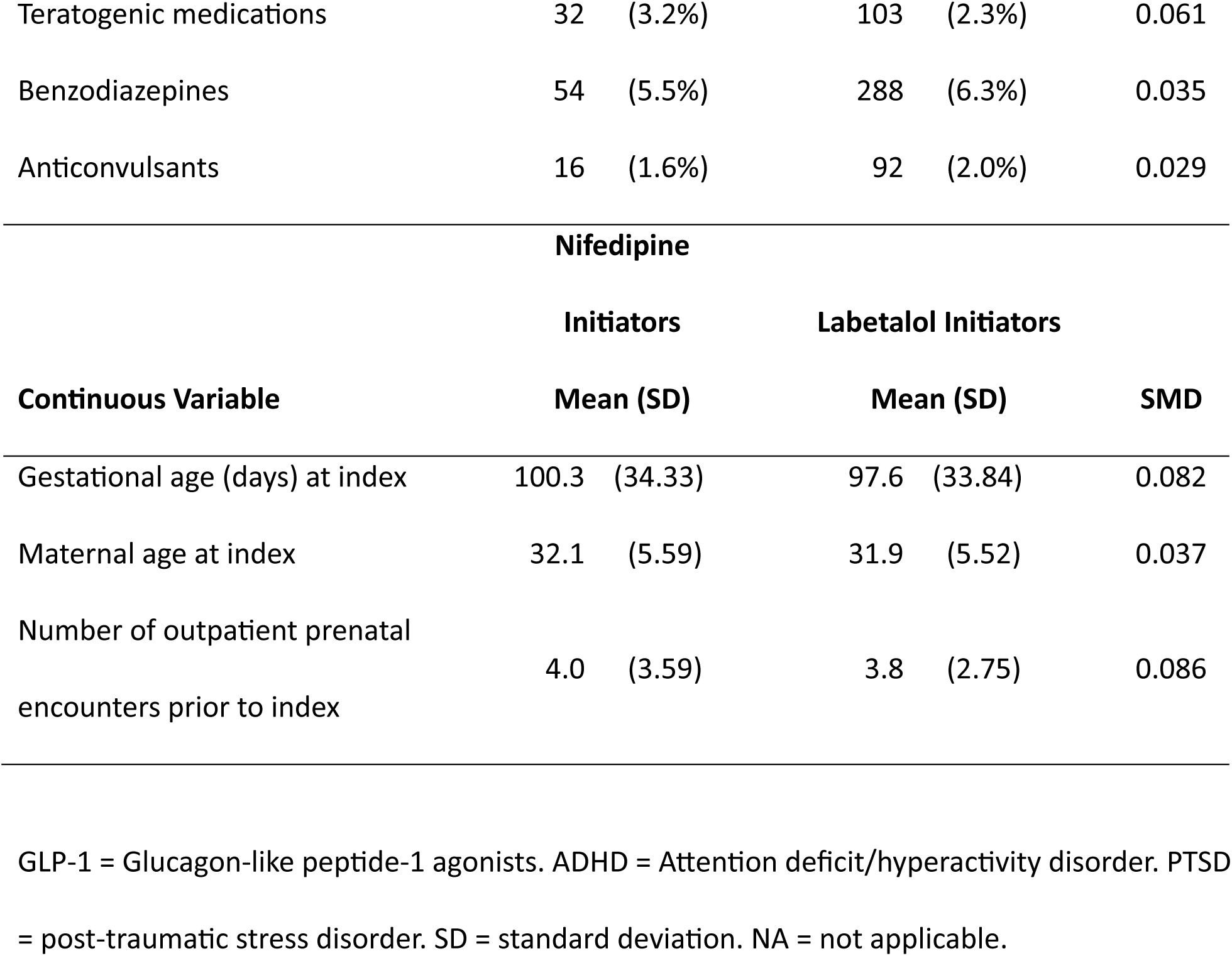
Descriptive statistics of the unweighted overall study population at baseline. Differences in proportions at each level of a covariate/characteristic are quantified via standardized mean differences (SMDs).

The treatment- and censoring-weighted absolute risk of preterm live birth among nifedipine initiators was 29.4% compared to 22.2% among labetalol initiators (Table 4, Figure 1, Figure S7). Nifedipine initiators thus had an estimated 7.2 per 100 pregnancies increased absolute risk (95% CI: 3.9, 10.5) of preterm live birth compared to labetalol initiators, corresponding to a RR of 1.32 (95% CI: 1.13, 1.55). The weighted risks of pregnancy loss were 7.0% versus 6.0%, respectively, with no indication of differences according to treatment group (RD: 1.1 per 100 pregnant people [95% CI: -0.7, 2.8]; RR: 1.18; [0.87, 1.59]). In *post hoc* analyses, the weighted absolute risk of preeclampsia among nifedipine initiators was 38.6% compared to 33.9% among labetalol initiators (RD: 4.7 per 100 pregnant people [95% CI: 0.7, 8.7], RR: 1.14 [95% CI: 1.01, 1.28]) (Table 4, Figure S8). Absolute risks of preeclampsia were lower when restricted to inpatient diagnoses (34.0% among nifedipine and 29.1% among labetalol initiators), but RDs and RRs were comparable.

**Figure 1.**
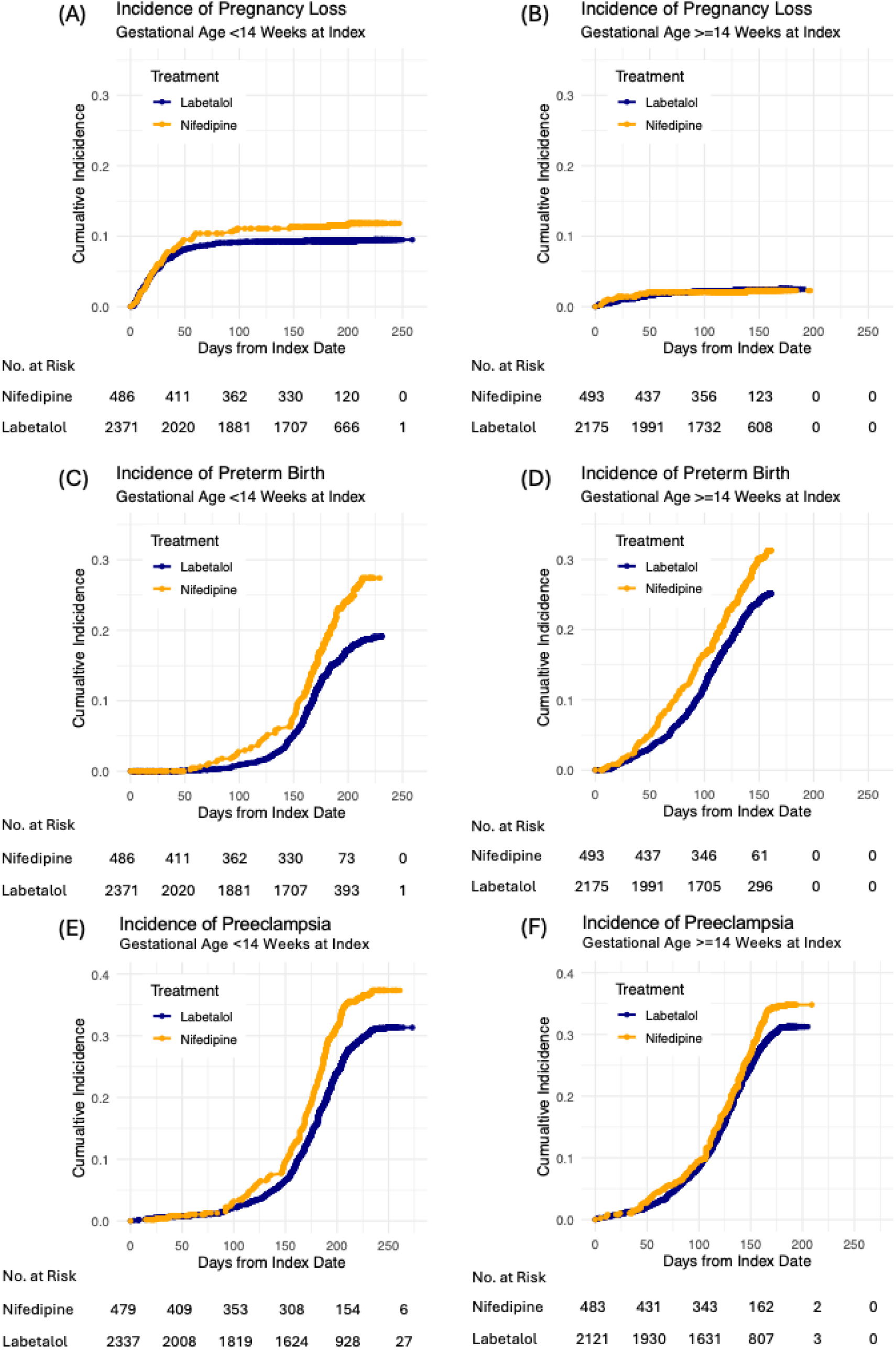
Cumulative incidence curves for pregnancy loss among nifedipine or labetalol initiators at (A) <14 and (B) ≥14 weeks’ gestation, for preterm live birth among nifedipine or labetalol initiators at (C) <14 and (D) ≥14 weeks’ gestation, and for preeclampsia (any service location) among nifedipine or labetalol initiators at (E) <14 and (F) ≥14 weeks’ gestation. Curves were estimated using a standardized mortality ratio and inverse probability of censoring weighted Aalen-Johansen estimator treating competing events as separate outcomes. Counts reflect the number of individuals in the at-risk population at that time, after implementing propensity score trimming.

**Table 4.**
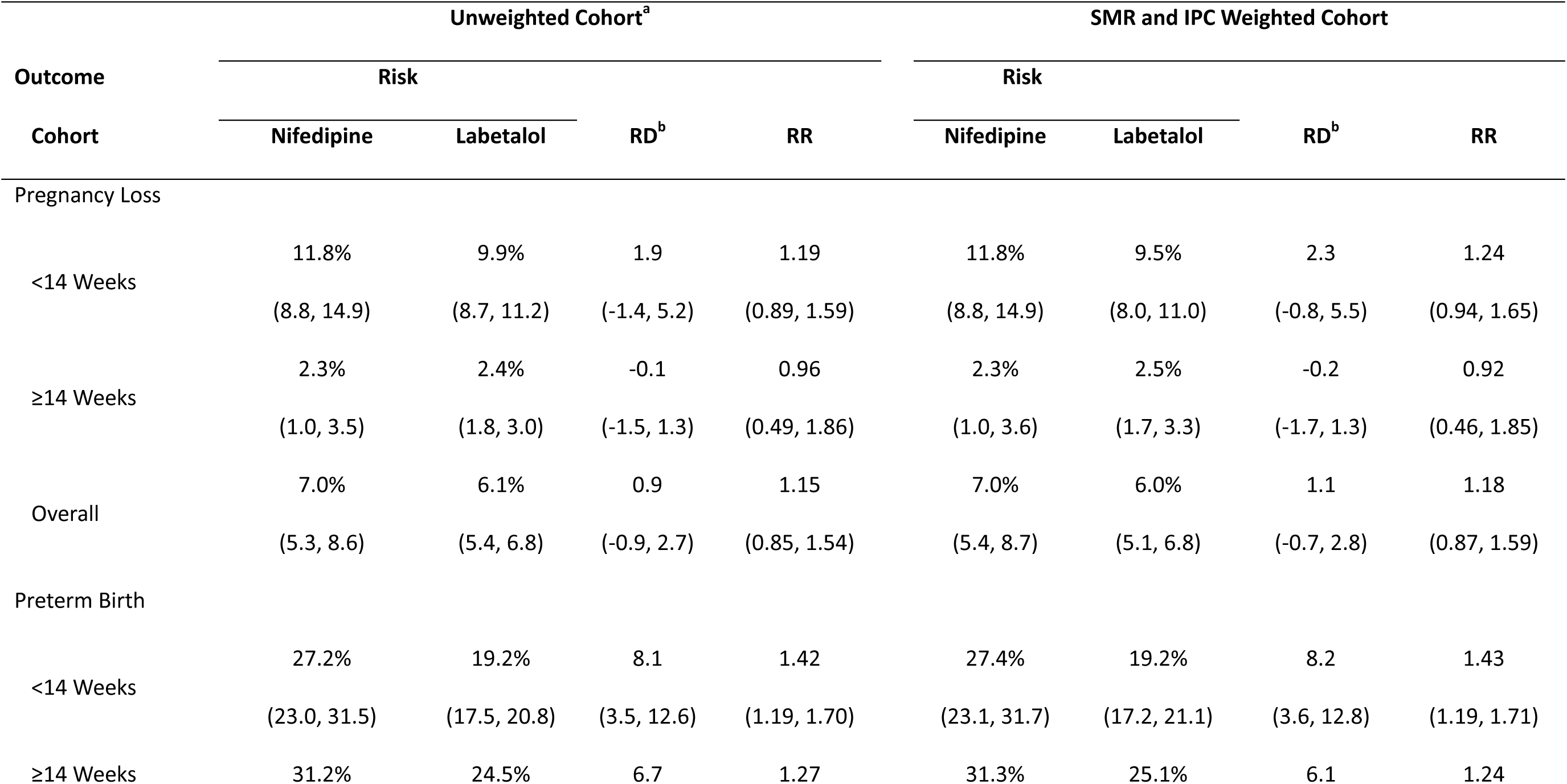
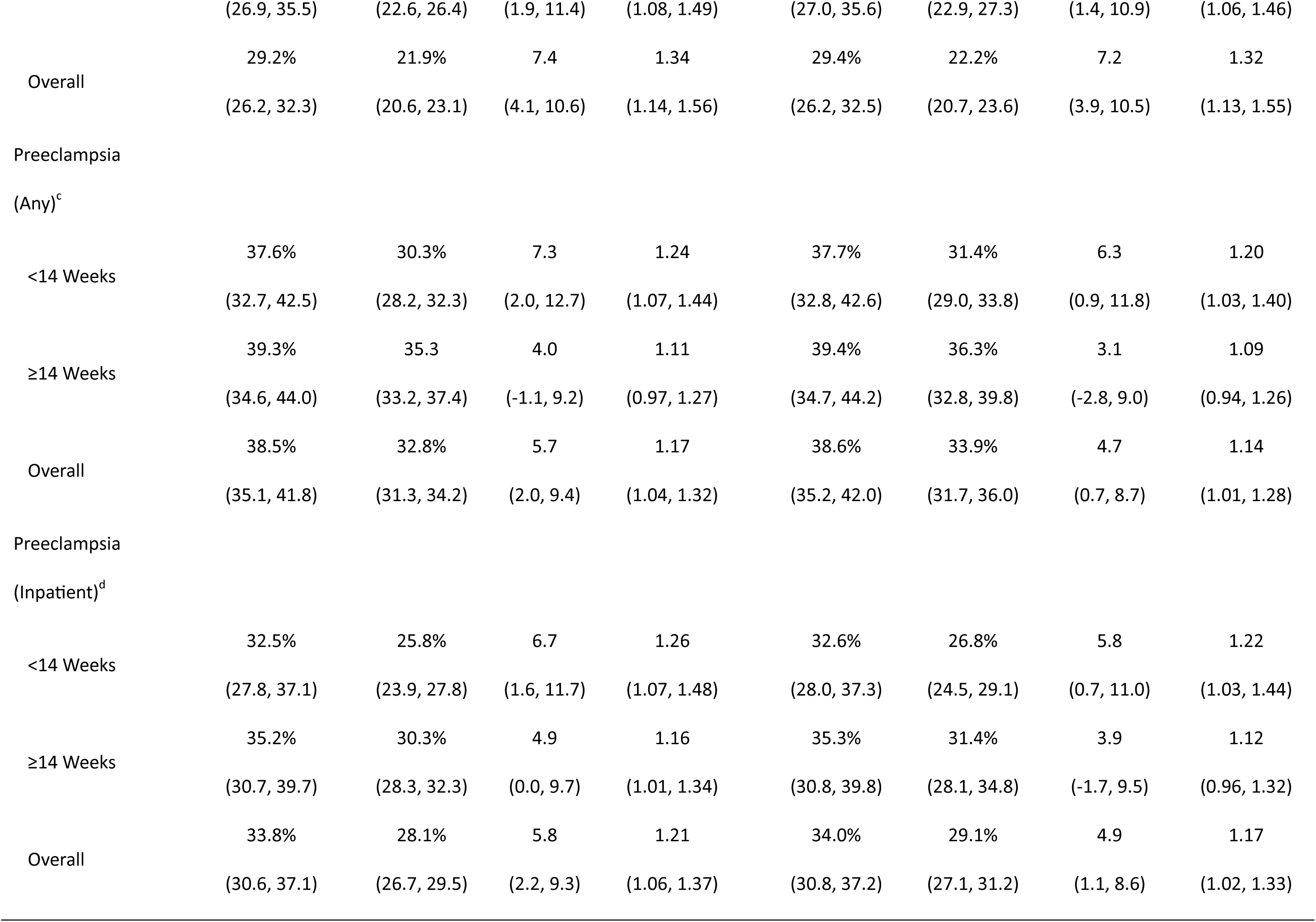
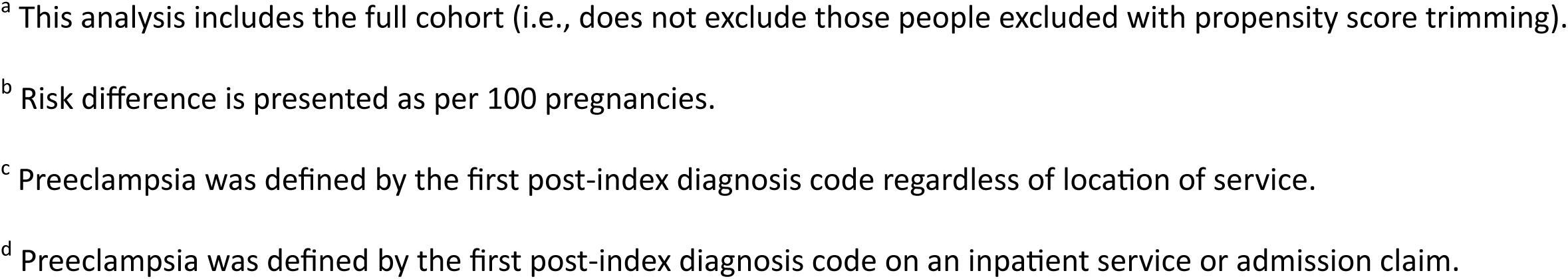
Risks, risk differences (RDs), and risk ratios (RRs) with 95% confidence intervals for pregnancy loss and preterm birth (<37 weeks) among the overall cohort and stratified by gestational age at initiation (<14 versus ≥14 weeks). Results include estimates among the unweighted cohort and the cohort applying both SMR and inverse probability of censoring (IPC) weights.

|  | Unweighted Cohort<sup>a</sup> |  |  |  | SMR and IPC Weighted Cohort |  |  |  |
| --- | --- | --- | --- | --- | --- | --- | --- | --- |
| Outcome | Risk |  |  |  | Risk |  |  |  |
| Cohort | Nifedipine | Labetalol | RD<sup>b</sup> | RR | Nifedipine | Labetalol | RD<sup>b</sup> | RR |
| Pregnancy Loss |  |  |  |  |  |  |  |  |
| <14 Weeks | 11.8% | 9.9% | 1.9 | 1.19 | 11.8% | 9.5% | 2.3 | 1.24 |
|  | (8.8, 14.9) | (8.7, 11.2) | (-1.4, 5.2) | (0.89, 1.59) | (8.8, 14.9) | (8.0, 11.0) | (-0.8, 5.5) | (0.94, 1.65) |
| ≥14 Weeks | 2.3% | 2.4% | -0.1 | 0.96 | 2.3% | 2.5% | -0.2 | 0.92 |
|  | (1.0, 3.5) | (1.8, 3.0) | (-1.5, 1.3) | (0.49, 1.86) | (1.0, 3.6) | (1.7, 3.3) | (-1.7, 1.3) | (0.46, 1.85) |
| Overall | 7.0% | 6.1% | 0.9 | 1.15 | 7.0% | 6.0% | 1.1 | 1.18 |
|  | (5.3, 8.6) | (5.4, 6.8) | (-0.9, 2.7) | (0.85, 1.54) | (5.4, 8.7) | (5.1, 6.8) | (-0.7, 2.8) | (0.87, 1.59) |
| Preterm Birth |  |  |  |  |  |  |  |  |
| <14 Weeks | 27.2% | 19.2% | 8.1 | 1.42 | 27.4% | 19.2% | 8.2 | 1.43 |
|  | (23.0, 31.5) | (17.5, 20.8) | (3.5, 12.6) | (1.19, 1.70) | (23.1, 31.7) | (17.2, 21.1) | (3.6, 12.8) | (1.19, 1.71) |
| ≥14 Weeks | 31.2% | 24.5% | 6.7 | 1.27 | 31.3% | 25.1% | 6.1 | 1.24 |
|  | (26.9, 35.5) | (22.6, 26.4) | (1.9, 11.4) | (1.08, 1.49) | (27.0, 35.6) | (22.9, 27.3) | (1.4, 10.9) | (1.06, 1.46) |
| Overall | 29.2% | 21.9% | 7.4 | 1.34 | 29.4% | 22.2% | 7.2 | 1.32 |
|  | (26.2, 32.3) | (20.6, 23.1) | (4.1, 10.6) | (1.14, 1.56) | (26.2, 32.5) | (20.7, 23.6) | (3.9, 10.5) | (1.13, 1.55) |
| Preeclampsia |  |  |  |  |  |  |  |  |
| (Any)<sup>c</sup> |  |  |  |  |  |  |  |  |
| <14 Weeks | 37.6% | 30.3% | 7.3 | 1.24 | 37.7% | 31.4% | 6.3 | 1.20 |
|  | (32.7, 42.5) | (28.2, 32.3) | (2.0, 12.7) | (1.07, 1.44) | (32.8, 42.6) | (29.0, 33.8) | (0.9, 11.8) | (1.03, 1.40) |
| ≥14 Weeks | 39.3% | 35.3 | 4.0 | 1.11 | 39.4% | 36.3% | 3.1 | 1.09 |
|  | (34.6, 44.0) | (33.2, 37.4) | (-1.1, 9.2) | (0.97, 1.27) | (34.7, 44.2) | (32.8, 39.8) | (-2.8, 9.0) | (0.94, 1.26) |
| Overall | 38.5% | 32.8% | 5.7 | 1.17 | 38.6% | 33.9% | 4.7 | 1.14 |
|  | (35.1, 41.8) | (31.3, 34.2) | (2.0, 9.4) | (1.04, 1.32) | (35.2, 42.0) | (31.7, 36.0) | (0.7, 8.7) | (1.01, 1.28) |
| Preeclampsia |  |  |  |  |  |  |  |  |
| (Inpatient)<sup>d</sup> |  |  |  |  |  |  |  |  |
| <14 Weeks | 32.5% | 25.8% | 6.7 | 1.26 | 32.6% | 26.8% | 5.8 | 1.22 |
|  | (27.8, 37.1) | (23.9, 27.8) | (1.6, 11.7) | (1.07, 1.48) | (28.0, 37.3) | (24.5, 29.1) | (0.7, 11.0) | (1.03, 1.44) |
| ≥14 Weeks | 35.2% | 30.3% | 4.9 | 1.16 | 35.3% | 31.4% | 3.9 | 1.12 |
|  | (30.7, 39.7) | (28.3, 32.3) | (0.0, 9.7) | (1.01, 1.34) | (30.8, 39.8) | (28.1, 34.8) | (-1.7, 9.5) | (0.96, 1.32) |
| Overall | 33.8% | 28.1% | 5.8 | 1.21 | 34.0% | 29.1% | 4.9 | 1.17 |
|  | (30.6, 37.1) | (26.7, 29.5) | (2.2, 9.3) | (1.06, 1.37) | (30.8, 37.2) | (27.1, 31.2) | (1.1, 8.6) | (1.02, 1.33) |

<sup>a</sup> This analysis includes the full cohort (i.e., does not exclude those people excluded with propensity score trimming).
<sup>b</sup> Risk difference is presented as per 100 pregnancies.
<sup>c</sup> Preeclampsia was defined by the first post-index diagnosis code regardless of location of service.
<sup>d</sup> Preeclampsia was defined by the first post-index diagnosis code on an inpatient service or admission claim.

RDs and RRs within gestational age strata aligned with overall estimates, though point estimates for preterm birth and preeclampsia were further from the null among initiators at <14 versus ≥14 weeks’ gestation. RDs and RRs from the secondary analyses were consistent with the primary results (Table S6).

### Pregnancy Outcomes Missing Not at Random

In preterm birth analyses, 13.2% of nifedipine and 12.5% of labetalol initiators were censored due to loss to follow-up; for pregnancy loss, these respective percentages were 13.3% and 12.6%; and for preeclampsia (any service location), 14.4% and 13.2%, respectively (Tables S7).

Full bounds on effect estimates were comparable to the primary study findings when unobserved outcomes were assumed the same across treatment but ranged across the null when unobserved outcomes differed by treatment (Figure 2, Figure S9, Table S8). For example, RD point estimates for preterm birth ranged from 6.0 to 6.7 per 100 for non-differential unobserved outcomes and -6.4 to 19.0 per 100 for differential unobserved outcomes

**Figure 2.**
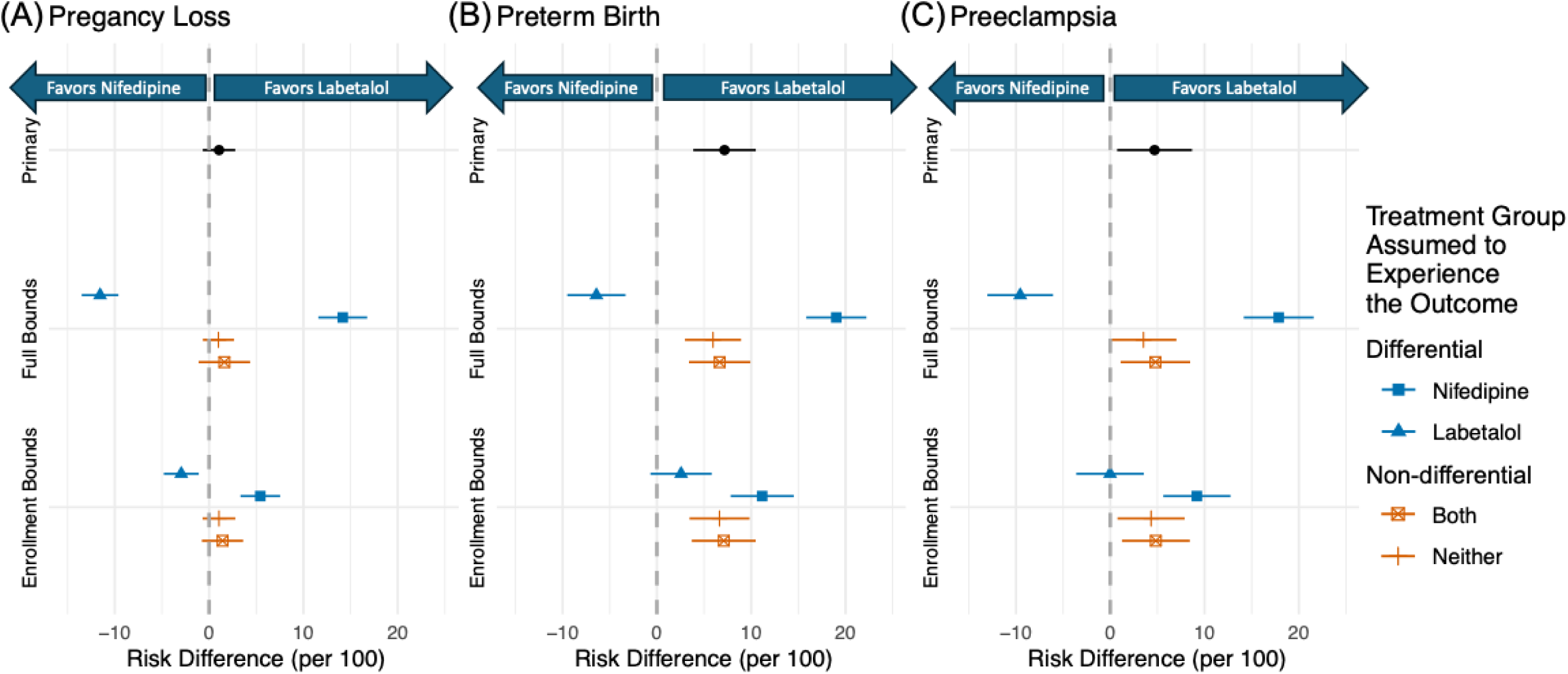
Risk differences (per 100 pregnancies) for (A) pregnancy loss, (B) preterm live birth (<37 weeks), and preeclampsia (any service location ≤14 days post-pregnancy outcome) from the primary analysis, full bounds, and enrollment-based bounds investigating pregnancy outcomes missing not at random. Within estimates from the bounds, color distinguishes whether the bound assumed that censoring for differential (blue) or non-differential (orange) by treatment. For full bounds, the shape indicates the treatment group of the censored pregnancies that we assumed experienced the outcome. For enrollment bounds, the shape indicates the group of censored pregnancies (censored >31 days after loss to follow-up) assumed to have a study outcome immediately at the start of the risk period.

After leveraging enrollment data, 31.5% of nifedipine and 29.6% of labetalol initiators with unobserved outcomes were assumed MNAR (i.e., disenrolled >31 days after censoring); for pregnancy loss, these percentages were 31.3% and 29.6%, respectively; and for preeclampsia (any service location), 30.9% and 30.4%, respectively (Tables S7). Pregnancies that disenrolled ≤31 days and >31 days after loss to follow-up had similar gestational ages at index and censoring (Figures S10-S11). The point estimates from the bounds corresponded to an increased risk of preterm birth among nifedipine versus labetalol initiators; over the different MNAR scenarios, the estimated RD ranged from 2.6 (95% CI: -0.7, 5.8) to 11.2 (95% CI: 7.8, 14.5) per 100 pregnancies. In contrast, the bounds for pregnancy loss (RD range: -3.0 [95% CI: -4.8, -1.1], 5.4 [95% CI: 3.3, 7.5]) and preeclampsia (any service location) (RD range: 0.0 [-3.6, 3.6], 9.2 [5.6, 12.8]) encompassed the null (Figure 2, Table S9). Bounds for preterm birth that included pregnancy loss were comparable to primary results (Table S9).

### Other Sensitivity Analyses

Complete case analyses were similar to primary analyses, though the risk of pregnancy loss was lower (Tables S10-S11). All other sensitivity analyses were consistent with the primary results (Tables S10-S13). Risks of pregnancy loss and preterm birth were higher among people with chronic hypertension diagnosis codes, but effect estimates were comparable.

### Descriptive Adherence Analysis

22.2% of nifedipine and 10.0% of labetalol initiators filled a prescription for a different antihypertensive than their initial treatment between their index and pregnancy outcome or loss to follow-up date. 41.7% of nifedipine and 44.2% of labetalol initiators had gaps between fills of 30 days or more, based on days’ supply. We found that such discontinuation could result in widely varying per-protocol effect estimates such that the bounds were wide and uninformative (Table S14).

## Discussion

We emulated a target trial comparing nifedipine and labetalol initiation before 23^0^ weeks’ gestation on the risk of pregnancy loss, preterm live birth, and preeclampsia among 5,557 pregnancies in a large database of commercially insured people in the US. We found that, compared to labetalol, nifedipine initiators had a higher adjusted risk of preterm birth, which may be partly explained by the observed increased risk of preeclampsia among nifedipine initiators. However, nifedipine and labetalol initiators experienced similar risks of pregnancy loss. Estimated treatment effects were similar for those who initiated treatment at earlier versus later gestational ages and robust to sensitivity analyses.

### Comparison with Previous Studies

Our findings for pregnancy loss are consistent with a secondary analysis of the CHAP trial, which found similar risks of fetal or neonatal death among nifedipine compared to labetalol users (RR=0.82 [95% CI: 0.41, 1.54]), and with a meta-analysis of trials investigating oral antihypertensives for treatments of hypertension in pregnancy (RR=1.32 [95% CI: 0.83, 2.13]) (Table S15).^7,8^ Similarly, our findings align with recent work among 4,122 deliveries in Kaiser Permanente’s electronic health record,^17^ which found that, compared to individuals who filled a prescription for labetalol before 36 weeks’ gestation, nifedipine users had similar odds of stillbirth or termination (odds ratio [OR]=1.10 [95% CI: 0.47, 2.01]).

Findings for preterm birth and preeclampsia, however, are less consistent. Our results align with those from a meta-analysis of trials for both preterm birth (RR=1.47 [95% CI: 1.11, 1.92]) and preeclampsia (RR=2.00 [95% CI: 1.15, 3.57]).^8^ In contrast, the CHAP reanalysis found no difference in preterm birth (RR=0.97 [95% CI: 0.80, 1.18]) or preeclampsia (RR=1.08 [95% CI: 0.88, 1.32]) risk by treatment. The Kaiser study found nifedipine associated with an increased risk of preterm birth (OR=1.25 [95% CI: 1.06, 1.46]) but not preeclampsia (OR=0.99 [95% CI: 0.84, 1.16]). Differences in the study designs (e.g., bias due to restricting to deliveries in the Kaiser data) and populations may explain apparently contrasting findings from the secondary CHAP analysis and Kaiser data for preterm birth and preeclampsia.

Several factors may lead clinicians to prescribe nifedipine over labetalol, resulting in confounded effect estimates. Providers may expect better adherence to once-daily nifedipine than twice- or thrice-daily labetalol,^58^ leading to preferential prescription of nifedipine to patients with higher risk of adverse outcomes. Further, calcium channel blockers like nifedipine are among recommended first-line antihypertensives for Black, non-pregnant adults but not others.^58,59^ This race-based guidance may lead providers to recommend nifedipine over labetalol to their Black pregnant patients, resulting in a higher proportion of Black people among nifedipine initiators, a group that also experiences a disproportionately high risk of preterm birth and preeclampsia.^60–62^ Despite using an active comparator to minimize differences between treatment groups,^63^ such prescribing patterns may introduce residual confounding by complex socioeconomic factors, race, or hypertension severity, which are unmeasured in MarketScan, resulting in the observed higher risk of preterm birth and preeclampsia in nifedipine versus labetalol initiatiors.^67^ Kaiser investigators also observed this increased risk of preterm birth with nifedipine, but adjusted for race, Hispanic ethnicity, and blood pressure, which was not possible in our analysis. Similar findings on preterm birth between our and the Kaiser study may suggest robustness to these unmeasured confounders. However, it is also possible that residual confounding in our data yielded biased estimates that coincidentally aligned with Kaiser’s more fully adjusted results. We believe this is less likely: differences in measured confounders between treatment groups were small, and the Kaiser study reported minimal differences by non-Hispanic, Black race and blood pressure, suggesting that these factors may not strongly influence treatment choice among pregnant people in real-world settings.

Differences in gestational ages at treatment decisions may cause differing treatment effects on preeclampsia between the studies. Abnormal placentation, a key pathway in preeclampsia development,^64^ is typically complete within the first 3 months of pregnancy,^65^ suggesting that the effect of medication choice on preeclampsia may be highly susceptible to timing of initiation. This appears supported by our results in which we observed weaker treatment effect estimates for preeclampsia among initiators at ≥14 weeks’ gestation versus <14 weeks’ gestation. Our study only included those who initiated nifedipine or labetalol before 23 weeks’ gestation, while the Kaiser study allowed treatment initiation, continuation, or switching up to 36 weeks with mean gestational ages at indexing fills of 20.8 and 18.8 weeks among nifedipine and labetalol users, respectively. Further, both the CHAP and Kaiser studies included prevalent users of antihypertensives pre-pregnancy (50.1% and 58.1% in CHAP and 58.3% and 61.5% in Kaiser, respectively), who may experience different effects on placentation than new users. Our study population does not include prevalent users nor late initiators, which prevents direct comparisons but suggests that these study findings may not be in conflict. Future studies are necessary to understand how these results may differ among prevalent users.

Adherence may also explain differing results from observational studies using administrative healthcare data versus CHAP data. Real-world adherence to nifedipine may be worse than labetalol due to its potential higher risk of adverse events, as seen in CHAP secondary analyses showing 35.7% of nifedipine versus 28.3% of labetalol users reported an adverse event (e.g., headache).^8^ In settings where adherence is less closely monitored than trials, this could contribute to worse outcomes among nifedipine initiators. While we found that similar percentages of pregnancies experienced gaps in their initiated medication across the treatment groups, more nifedipine than labetalol initiators filled a prescription for a different antihypertensive over follow-up, which suggests nifedipine users may have experienced more side effects, inadequate blood pressure control, or both. CHAP’s rigorous adherence protocols may have resulted in intention to treat estimates approximating per-protocol effects, while our results may be explained by differing adherence across initiators. It may be reasonable, then, to expect the comparative effect of nifedipine versus labetalol to differ among real-world pregnant populations.

As with all studies using insurance claims to study pregnancy, gestational age of pregnancy was estimated based on recorded codes, which is likely to result in measurement error,^66^ particularly for pregnancies ending in outcomes other than live birth. Such measurement error could result in a misidentification of our target population (e.g., including individuals with gestational instead of chronic hypertension) as well as misclassification of covariates (e.g., pregestational diabetes) and outcomes (e.g., preterm birth). While we cannot rule out residual bias,^40^ sensitivity analyses suggest that our results were robust to realistic mis-estimation of gestational age.

Our study also has important strengths that support the robustness of our results among real-world populations. We focused on antihypertensive initiation, and while questions regarding treatment decisions among prevalent users are important, our approach avoids prevalent-user bias.^42,67,68^ Combining new and prevalent users in analyses may bias effect estimates of prenatal antihypertensive use because the decision to continue, switch, or discontinue antihypertensives likely shares common causes with our study outcomes (e.g., hypertension severity).^24^ We also reduced immortal time bias by requiring pregnancies be observed before initiation.^36^

Additionally, we included pregnancies with unobserved outcomes. While this introduced challenges due to the potential for informative missingness, our findings were robust across certain assumptions regarding the treatment-specific outcome missingness mechanism.^13^ Though our conclusions could change in scenarios where the true unobserved outcome mechanisms differ substantially between nifedipine and labetalol initiators, we are unable to identify plausible scenarios for which this could occur.

## Conclusions

Among pregnant people initiating antihypertensive treatment before 23^0^ weeks’ gestation, compared to labetalol users, nifedipine users may be at greater risk of preterm birth and preeclampsia but not pregnancy loss. While high quality evidence from trials shows a clear treatment benefit for antihypertensive therapy with either nifedipine or labetalol compared to no treatment,^1,4,5^ our results offer valuable evidence to support patient-provider decisions between initiating the two medications for chronic hypertension in pregnancy. Future work should evaluate the effects of changes to antihypertensive treatment regimens among pre-pregnancy users of antihypertensives, methods for improving medication adherence, and potential effect modification within patient subgroups.

## Data Availability

Data included in these analyses can be obtained through a data use agreement with Merative. Copyright ©2025 Merative. All Rights Reserved. SAS code and code lists used to derive a cohort of pregnancies from MarketScan claims are available on GitHub (https://github.com/Safe2Treat/Pregnancy-algorithm-prospective). SAS and R code used to finalize and analyze the cohort data are also available on GitHub (https://github.com/chasedlatour/Nifedipine_v_Labetalol_MarketScan).

## Acknowledgements

We would like to thank Virginia Pate for code and advice on using MarketScan claims data and implementing these analyses within the full data source; Stephen Cole, PhD, for his advice on implementing the Aalen-Johansen estimator in combination with our proposed sensitivity analyses for informative censoring; and Paul Zivich, PhD, for sharing his and colleagues’ work-in-progress describing bounds on the per-protocol effect estimate for time-to-event outcomes. ChatGPT was used to assist with code de-bugging and to improve grammar and clarity in a few sections of the final manuscript draft (OpenAI, San Francisco, California).

## References

1. The American College of Obstetricians and Gynecologists. ACOG Practice Bulletin: Chronic Hypertension in Pregnancy. Obstetrics & Gynecology. 2019;133(76):168–186.

2. Bramham K, Parnell B, Nelson-Piercy C, Seed PT, Poston L, Chappell LC. Chronic hypertension and pregnancy outcomes: Systematic review and meta-analysis. BMJ. 2014;348:g2301. doi:10.1136/bmj.g2301

3. Leonard SA, Siadat S, Main EK, et al. Chronic Hypertension During Pregnancy: Prevalence and Treatment in the United States, 2008–2021. Hypertension. Published online June 17, 2024. doi:10.1161/HYPERTENSIONAHA.124.22731

4. Tita AT, Szychowski JM, Boggess K, et al. Treatment for Mild Chronic Hypertension during Pregnancy. New England Journal of Medicine. Published online April 2, 2022. doi:10.1056/NEJMoa2201295

5. Society for Maternal-Fetal Medicine and PC. Society for Maternal-Fetal Medicine Statement: Antihypertensive therapy for mild chronic hypertension in pregnancy–The Chronic Hypertension and Pregnancy trial. Am J Obstet Gynecol. 2022;227(2):B24–B27. doi:10.1016/j.ajog.2022.04.011

6. American College of Obstetricians and Gynecologists’ Committee on Clinical Practice Guidelines. Clinical Guidance for the Integration of the Findings of the Chronic Hypertension and Pregnancy (CHAP) Study. Practice Advisory. March 2024. Accessed May 26, 2024. https://www.acog.org/clinical/clinical-guidance/practice-advisory/articles/2022/04/clinical-guidance-for-the-integration-of-the-findings-of-the-chronic-hypertension-and-pregnancy-chap-study

7. Hup RJ, Damen JAA, Terstappen J, et al. Oral antihypertensive treatment during pregnancy: a systematic review and network meta-analysis. Am J Obstet Gynecol. Published online April 2025. doi:10.1016/j.ajog.2025.04.011

8. Sanusi AA, Leach J, Boggess K, et al. Pregnancy Outcomes of Nifedipine Compared With Labetalol for Oral Treatment of Mild Chronic Hypertension. Obstetrics & Gynecology. Published online July 23, 2024. doi:10.1097/aog.0000000000005613

9. Ashworth D, Battersby C, Green M, et al. Which antihypertensive treatment is better for mild to moderate hypertension in pregnancy? BMJ. Published online January 18, 2022:e066333. doi:10.1136/bmj-2021-066333

10. Tan YY, Papez V, Chang WH, Mueller SH, Denaxas S, Lai AG. Comparing clinical trial population representativeness to real-world populations: an external validity analysis encompassing 43 895 trials and 5 685 738 individuals across 989 unique drugs and 286 conditions in England. Lancet Healthy Longev. 2022;3(10):e674–e689. doi:10.1016/S2666-7568(22)00186-6

11. Westreich D, Edwards JK. Invited Commentary: Every Good Randomization Deserves Observation. Am J Epidemiol. 2015;182(10):857–860. doi:10.1093/aje/kwv200

12. DiCesare E, Huybrechts KF, Bateman BT, Lii J, Straub L. Antihypertensive Treatment Adherence during Pregnancy by Race and Ethnicity. Am J Obstet Gynecol. Published online May 22, 2025. doi:10.1016/j.ajog.2025.05.015

13. Latour CD, Edwards JK, Jonsson Funk M, Suarez EA, Boggess K, Wood ME. Bias in studies of prenatal exposures using real-world data due to pregnancy identification method. ArXiv. Published online 2025. 10.48550/arXiv.2504.12415

14. Suarez EA, Landi SN, Conover MM, Jonsson Funk M. Bias from restricting to live births when estimating effects of prescription drug use on pregnancy complications: A simulation. Pharmacoepidemiol Drug Saf. 2018;27(3):307–314. doi:10.1002/pds.4387

15. Liew Z, Olsen J, Cui X, Ritz B, Arah OA. Bias from conditioning on live birth in pregnancy cohorts: An illustration based on neurodevelopment in children after prenatal exposure to organic pollutants. Int J Epidemiol. 2015;44(1):345–354. doi:10.1093/ije/dyu249

16. Leung M, Kioumourtzoglou MA, Raz R, Weisskopf MG. Bias due to selection on live births in studies of environmental exposures during pregnancy: A simulation study. Environ Health Perspect. 2021;129(4). doi:10.1289/EHP7961

17. Dublin S, Idu A, Avalos LA, et al. Maternal and neonatal outcomes of antihypertensive treatment in pregnancy: A retrospective cohort study. PLoS One. 2022;17(5):e0268284. doi:10.1371/journal.pone.0268284

18. Smith N, Kwon Kim S, Goyert G, Lin CH, Rose C, Pitts DS. Nifedipine outperforms labetalol: A comparative analysis of hypertension management in black pregnancies. Pregnancy Hypertens. 2024;37. doi:10.1016/j.preghy.2024.101147

19. Hernández-Díaz S, Huybrechts KF, Chiu YH, et al. Emulating a target trial of interventions initiated during pregnancy with healthcare databases: the example of COVID-19 vaccination. Epidemiology. 2023;34:238–246. doi:10.1097/EDE.0000000000001562

20. Hernán MA, Sauer BC, Hernández-Díaz S, Platt R, Shrier I. Specifying a target trial prevents immortal time bias and other self-inflicted injuries in observational analyses. J Clin Epidemiol. 2016;79:70–75. doi:10.1016/j.jclinepi.2016.04.014

21. Hernán MA, Wang W, Leaf DE. Target Trial Emulation: A Framework for Causal Inference from Observational Data. JAMA. 2022;328(24):2446–2447. doi:10.1001/jama.2022.21383

22. Hernán MA, Dahabreh IJ, Dickerman BA, Swanson SA. The Target Trial Framework for Causal Inference From Observational Data: Why and When Is It Helpful? Ann Intern Med. Published online February 18, 2025. doi:10.7326/ANNALS-24-01871

23. Margulis A V, Huybrechts K. Identification of pregnancies in healthcare data: A changing landscape. Pharmacoepidemiol Drug Saf. 2023;32(1):84–86. doi:10.1002/pds.5526

24. Huybrechts KF, Bateman BT, Hernández-Díaz S. Use of real-world evidence from healthcare utilization data to evaluate drug safety during pregnancy. Pharmacoepidemiol Drug Saf. 2019;28(7):906–922. doi:10.1002/pds.4789

25. Suarez EA, Boggess K, Engel SM, Stürmer T, Lund JL, Funk MJ. Ondansetron use in early pregnancy and the risk of late pregnancy outcomes. Pharmacoepidemiol Drug Saf. 2021;30(2):114–125. doi:10.1002/pds.5151

26. Ailes EC, Simeone RM, Dawson AL, Petersen EE, Gilboa SM. Using insurance claims data to identify and estimate critical periods in pregnancy: An application to antidepressants. Birth Defects Res A Clin Mol Teratol. 2016;106(11):927–934. doi:10.1002/bdra.23573

27. Ailes EC, Zhu W, Clark EA, et al. Identification of pregnancies and their outcomes in healthcare claims data, 2008-2019: An algorithm. PLoS One. 2023;18(4):e0284893. doi:10.1371/journal.pone.0284893

28. Moll K, Wong HL, Fingar K, et al. Validating Claims-Based Algorithms Determining Pregnancy Outcomes and Gestational Age Using a Linked Claims-Electronic Medical Record Database. Drug Saf. 2021;44(11):1151–1164. doi:10.1007/s40264-021-01113-8

29. Sarayani A, Wang X, Thai TN, Albogami Y, Jeon N, Winterstein AG. Impact of the transition from ICD–9–CM to ICD–10–CM on the identification of pregnancy episodes in us health insurance claims data. Clin Epidemiol. 2020;12:1129–1138. doi:10.2147/CLEP.S269400

30. MacDonald SC, Cohen JM, Panchaud A, McElrath TF, Huybrechts KF, Hernández-Díaz S. Identifying pregnancies in insurance claims data: Methods and application to retinoid teratogenic surveillance. Pharmacoepidemiol Drug Saf. 2019;28(9):1211–1221. doi:10.1002/pds.4794

31. Andrade SE, Shinde M, Moore Simas TA, et al. Validation of an ICD-10-based algorithm to identify stillbirth in the Sentinel System. Pharmacoepidemiol Drug Saf. 2021;30(9):1175–1183. doi:10.1002/pds.5300

32. Bertoia ML, Phiri K, Clifford CR, et al. Identification of pregnancies and infants within a US commercial healthcare administrative claims database. Pharmacoepidemiol Drug Saf. 2022;31(8):863–874. doi:10.1002/pds.5483

33. Suarez E. Sentinel Pregnancy Tool and Transition to ICD-10 [slide presentation]. Published online 2019.

34. Suarez EA, Nguyen M, Zhang D, et al. Novel methods for pregnancy drug safety surveillance in the FDA Sentinel System. Pharmacoepidemiol Drug Saf. Published online 2022. doi:10.1002/pds.5512

35. Ayoola AB, Nettleman MD, Stommel M, Canady RB. Time of pregnancy recognition and prenatal care use: A population-based study in the United States. Birth. 2010;37(1):37–43. doi:10.1111/j.1523-536X.2009.00376.x

36. Wood ME, Latour CD, Petito LC. Treatments for Pregestational Chronic Conditions during Pregnancy: Emulating a Target Trial with a Treatment Decision Design.; 2023.

37. Chen L, Shortreed SM, Easterling T, et al. Identifying Hypertension in Pregnancy using Electronic Medical Records: The importance of Blood Pressure Values. Pregnancy Hypertens. 2020;19:112–118. doi:10.1016/j.preghy.2020.01.001

38. Shrier I, Platt RW. Reducing bias through directed acyclic graphs. BMC Med Res Methodol. 2008;8(70). doi:10.1186/1471-2288-8-70

39. Howe CJ, Bailey ZD, Raifman JR, Jackson JW. Recommendations for Using Causal Diagrams to Study Racial Health Disparities. Am J Epidemiol. Published online August 2, 2022. doi:10.1093/aje/kwac140

40. Daniel RM, Kenward MG, Cousens SN, De Stavola BL. Using causal diagrams to guide analysis in missing data problems. Stat Methods Med Res. 2012;21(3):243–256. doi:10.1177/0962280210394469

41. Howe CJ, Cole SR, Lau B, Napravnik S, Eron JJ. Selection Bias Due to Loss to Follow Up in Cohort Studies. Epidemiology. 2016;27(1):91–97. doi:10.1097/EDE.0000000000000409

42. Lund JL, Richardson DB, Stürmer T. The Active Comparator, New User Study Design in Pharmacoepidemiology: Historical Foundations and Contemporary Application. Curr Epidemiol Rep. 2015;2(4):221–228. doi:10.1007/s40471-015-0053-5

43. Stürmer T, Wyss R, Glynn RJ, Brookhart MA. Propensity scores for confounder adjustment when assessing the effects of medical interventions using nonexperimental study designs. J Intern Med. 2014;275(6):570–580. doi:10.1111/joim.12197

44. Brookhart MA, Wyss R, Layton JB, Stürmer T. Propensity Score Methods for Confounding Control in Non-Experimental Research. Circ Cardiovasc Qual Outcomes. 2013;6(5):604–611. doi:10.1161/CIRCOUTCOMES.113.000359

45. Aalen OO, Johansen S. An Empirical Transition Matrix for Non-Homogeneous Markov Chains Based on Censored Observations. Scandinavian Journal of Statistics. 1978;5(3):141–150. https://www.jstor.org/stable/4615704?seq=1&cid=pdf-

46. Bluhmki T, Fietz AK, Stegherr R, et al. Multistate methodology improves risk assessment under time-varying drug intake—a new view on pregnancy outcomes following coumarin exposure. Pharmacoepidemiol Drug Saf. 2019;28(5):616–624. doi:10.1002/pds.4710

47. Latour CD, Klose M, Edwards JK, Song Z, Jonsson Funk M, Wood ME. Healthy Live Births as Censoring Versus Competing Events in Studies of Prenatal Medication Use. Paediatr Perinat Epidemiol. Published online July 7, 2025. doi:10.1111/ppe.70043

48. Efron B. Bootstrap Methods: Another Look at the Jackknife. In: Kotz S, Johnson NL, eds. Breakthroughs in Statistics. Springer Series in Statistics. Springer; 1992:569–593. doi:10.1007/978-1-4612-4380-9_41

49. WHO Recommendations on Drug Treatment for Non-Severe Hypertension in Pregnancy.; 2020. Accessed March 29, 2025. https://www.who.int/publications/i/item/9789240008793

50. American College of Obstetricians and Gynecologists’ Committee on Practice Bulletins-Obstetrics. ACOG Practice Bulletin No. 222: Gestational Hypertension and Preeclampsia. Obstetrics & Gynecology. 2020;135(6):e237–e260. doi:10.1097/AOG.0000000000003891

51. Palmsten K, Huybrechts KF, Kowal MK, Mogun H, Hernández-Díaz S. Validity of maternal and infant outcomes within nationwide Medicaid data. Pharmacoepidemiol Drug Saf. 2014;23(6):646–655. doi:10.1002/pds.3627

52. Chomistek AK, Phiri K, Doherty MC, et al. Development and Validation of ICD-10-CM-based Algorithms for Date of Last Menstrual Period, Pregnancy Outcomes, and Infant Outcomes. Drug Saf. 2023;46(2):209–222. doi:10.1007/s40264-022-01261-5

53. Schisterman EF, DeVilbiss EA, Perkins NJ. A method to visualize a complete sensitivity analysis for loss to follow-up in clinical trials. Contemp Clin Trials Commun. 2020;19. doi:10.1016/j.conctc.2020.100586

54. Cole SR, Hudgens MG, Edwards JK, et al. Nonparametric Bounds for the Risk Function. Am J Epidemiol. 2019;188(4):632–636. doi:10.1093/aje/kwz013

55. Groenwold RHH, Palmer TM, Tilling K. To Adjust or Not to Adjust? When a “Confounder” Is Only Measured After Exposure. Epidemiology. 2021;32(2):194–201. doi:10.1097/EDE.0000000000001312

56. Murray EJ, Caniglia EC, Petito LC. Causal survival analysis: A guide to estimating intention-to-treat and per-protocol effects from randomized clinical trials with non-adherence. Research Methods in Medicine & Health Sciences. 2021;2(1):39–49. doi:10.1177/2632084320961043

57. Swanson SA, Holme Ø, Løberg M, et al. Bounding the per-protocol effect in randomized trials: an application to colorectal cancer screening. Trials. 2015;16(1):541. doi:10.1186/s13063-015-1056-8

58. Whelton PK, Carey RM, Aronow WS, et al. 2017 ACC/AHA/AAPA/ABC/ACPM/AGS/APhA/ASH/ASPC/NMA/PCNA Guideline for the Prevention, Detection, Evaluation, and Management of High Blood Pressure in Adults: A Report of the American College of Cardiology/American Heart Association Task Force on Clinical Practice Guidelines. J Am Coll Cardiol. 2018;71(19):e127-e248. doi:10.1016/j.jacc.2017.11.006

59. Peter JG, Ntusi NAB, Ntsekhe M. Are Recommendations That Favor Other Agents Over Angiotensin-Converting Enzyme Inhibitors in Africans With Hypertension Justified? Circulation. 2024;149(11):804–806. doi:10.1161/CIRCULATIONAHA.123.065887

60. Braveman P, Dominguez TP, Burke W, et al. Explaining the Black-White Disparity in Preterm Birth: A Consensus Statement From a Multi-Disciplinary Scientific Work Group Convened by the March of Dimes. Frontiers in Reproductive Health. 2021;3. doi:10.3389/frph.2021.684207

61. Atkinson JA, Carmichael SL, Leonard SA. Hypertensive Disorders in Pregnancy: Differences by Hispanic Ethnicity and Black Race. J Racial Ethn Health Disparities. Published online November 5, 2024. doi:10.1007/s40615-024-02224-5

62. Ford ND, Cox S, Ko JY, et al. Hypertensive Disorders in Pregnancy and Mortality at Delivery Hospitalization-United States, 2017-2019. Morbidity and Mortality Weekly Report. 2022;71:585–591.

63. Sendor R, Stürmer T. Core concepts in pharmacoepidemiology: Confounding by indication and the role of active comparators. Pharmacoepidemiol Drug Saf. 2022;31(3):261–269. doi:10.1002/pds.5407

64. Friedman AM. Placentation, Hypertensive Disorders, and Neonatal Outcomes. Paediatr Perinat Epidemiol. Published online February 17, 2025. doi:10.1111/ppe.70006

65. Herrick EJ, Bordoni B. Embryology, Placenta. In: StatPearls [Internet]. StatPearls Pbulishing; 2025. Accessed March 31, 2025. https://www.ncbi.nlm.nih.gov/books/NBK551634

66. Chiu YH, Huybrechts KF, Zhu Y, et al. Internal validation of gestational age estimation algorithms in health-care databases using pregnancies conceived through fertility procedures. Am J Epidemiol. Published online April 6, 2024. doi:10.1093/aje/kwae045

67. Ray WA. Evaluating Medication Effects Outside of Clinical Trials: New-User Designs. Am J Epidemiol. 2003;158(9):915–920. doi:10.1093/aje/kwg231

68. Olawore O, Stürmer T, Glynn RJ, Lund JL. The healthy user effect in pharmacoepidemiology. Am J Epidemiol. 2025;194(7):2023–2029. doi:10.1093/aje/kwae358

